# Coach-BP: Computer Vision-Based Assessment and Correction of Blood Pressure Measurement Technique

**DOI:** 10.64898/2026.10.29.26366628

**Authors:** Rahul Biswas, Alvin Joseph Abad, Jesuit Nugas, Joan Shim, Mark J Pletcher, Julio A Lamprea-Montealegre, Reza Abbasi-Asl

## Abstract

**Background:** Accurate blood pressure (BP) measurement, essential for hypertension care, requires proper technique. Posture errors can raise systolic BP by more than 20 mm Hg, yet technique is rarely verified objectively. We aimed to develop and evaluate Coach-BP, a computer vision system that detects technique errors from video to support real-time coaching.

**Methods:** We enrolled adults attending an academic cardiovascular prevention center between February and April 2026 and filmed BP measurements with front and side cameras. We analyzed 408 encounters from 225 participants. Coach-BP uses machine learning models trained on clinician labels to assess five American Heart Association criteria using pose, visual context, and audio-visual features. We evaluated its accuracy against expert clinician ratings by leave-one-participant-out cross-validation. We then examined robustness to simulated camera artifacts and interpreted the features important for each criterion.

**Findings:** At baseline, 138 (62%) of 223 participants had at least one technique error and 54 (24%) two or more. Coach-BP achieved a mean area under the receiver operating characteristic curve (AUROC) of 0·93 (95% CI 0·90–0·94), with 0·97 (0·94–0·99) for legs uncrossed, 0·97 (0·93–1·00) for no talking, 0·92 (0·85–0·98) for back supported, 0·91 (0·86–0·95) for feet flat, and 0·86 (0·81–0·91) for arm supported at heart level. Mean balanced accuracy was 0·88 (0·85–0·90). Performance was robust to simulated camera shift, zoom, and rotation. Its models relied on features clinicians would assess, such as cuffed-hand height and foot position. One model classified back support similarly from either camera.

**Interpretation:** Coach-BP accurately detected errors in the five guideline-recommended criteria of BP measurement technique from clinic video, enabling objective assessment of each criterion.

**Funding:** University of California San Francisco Cardiology Scholars Award.

**Research in context:** *Evidence before this study:* We searched PubMed and Europe PMC from database inception to Oct 10, 2026, without language restriction, for studies in adults that automatically assessed any guideline-recommended patient condition during seated upper-arm blood pressure measurement, using terms including “blood pressure”, “measurement technique”, “posture”, “computer vision”, “pose estimation”, and “sensor”. A systematic review of 328 studies and a randomized crossover trial of arm position have quantified the effect of individual errors, several of which raise systolic blood pressure by 5 mm Hg or more, and studies that observed or filmed patients measuring their blood pressure found errors in almost all patients. Three studies assessed measurement conditions automatically, all with the same sensor-equipped chair, which recorded rest time, back support, crossed legs, and ambient noise during self-measurement. They were small (41–113 participants), and only one compared the chair’s output with direct observation. In one, adherence was high only for the conditions on which the chair gave live guidance. None used video, and the chair did not assess arm or foot position. The evidence on the effect of errors includes a randomized trial, whereas the evidence on automated assessment is limited to these small observational studies of one device.

*Added value of this study:* In 225 adults contributing 408 blood pressure measurement encounters, technique errors were present at the first reading in 138 (62%) of 223 participants and did not differ significantly across the self-reported characteristics examined, including home blood pressure monitoring. Among 171 participants re-measured after staff-delivered coaching, the number with any technique error fell from 128 (75%) to 12 (7%). Coach-BP assessed all five guideline-recommended posture and behavior criteria directly from clinic video, with a mean area under the receiver operating characteristic curve (AUROC) of 0·93 under leave-one-participant-out cross-validation. Coach-BP matched or improved on rule-based baselines for all five criteria, with gains for feet flat, the arm criterion, back supported, and no talking. Performance remained stable when the image was shifted, rescaled, or rotated, suggesting robustness to differences in how the camera frames the patient. One back-support model performed similarly from the side and front cameras, so a single front camera placed as in the study could assess all five criteria.

*Implications of all the available evidence:* Technique errors were common, including among patients who already monitor blood pressure at home, and most were corrected after brief staff coaching. Coach-BP shows that five criteria of blood pressure measurement technique can be assessed automatically and accurately from ordinary video. Together, these findings support automated feedback at the time of measurement as a scalable way to improve technique, particularly at home, where no one checks it. Prospective studies should now test this in home settings and more diverse populations.

## Introduction

Hypertension is a leading modifiable contributor to cardiovascular morbidity and mortality, affecting more than 1 billion adults worldwide.^1–3^ Yet only about one in five people with hypertension have their blood pressure controlled.^3^ Despite the importance of accurate blood pressure (BP) measurement for clinical decision-making, measurements are frequently obtained under conditions that introduce substantial error.

The American Heart Association (AHA) and other professional societies recommend that patients be seated with their back supported, feet flat on the floor, legs uncrossed, without talking, and the measurement arm supported at heart level.^4–6^ Deviations from these recommendations are common^4^ and raise measured systolic BP,^7^ in some studies by more than 20 mm Hg.^8^ Errors of this size can lead to hypertension being misdiagnosed and treatment being increased unnecessarily.^7,9^

Despite guideline recommendations, BP measurement technique is rarely verified in routine practice. Office measurement is often poorly standardized,^9^ the errors behind a recorded reading can rarely be identified afterwards,^8^ and checking technique would require an observer at every reading. Automated office devices, which take several unattended readings, reduce observer-related error but do not verify posture.^10^ Home monitoring, which enables longitudinal assessment and timely titration,^9,11^ moves measurements used for treatment decisions to unsupervised settings where technique is unverified.

Computer vision may make automated verification of technique feasible.^12^ Here we present Coach-BP, a computer vision system intended to check, at the time of a seated measurement, whether each of five technique criteria is met, so that technique can be corrected before a reading is used for clinical decisions (Figure 1). Coach-BP uses pose-estimation, visual-representation, and audio-visual models to assess body positioning and speech from standard video recordings.^13–16^ In this study, we evaluated its accuracy for the five recommended criteria from front and side video of adults at a cardiovascular prevention clinic.

**Figure 1:**
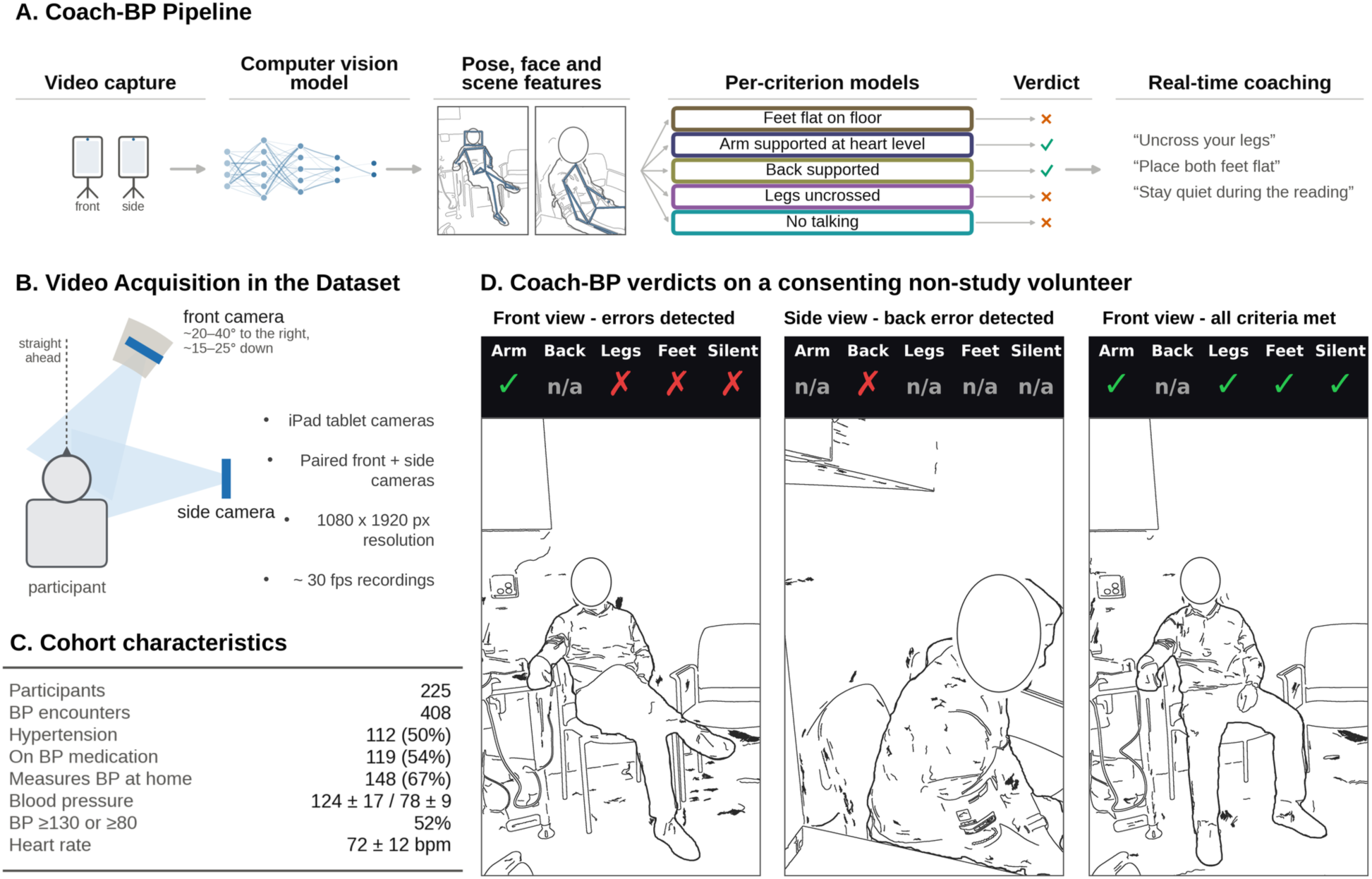
Overview of Coach-BP. (A) Processing pipeline. Front and side videos are processed by pose, face, and scene models, and the cuffed arm is located with an open-vocabulary object detector. Their outputs feed one model for each American Heart Association criterion, which returns a verdict. The final panel shows the feedback Coach-BP is intended to give. (B) Video acquisition, shown from above. Front and side tablet cameras recorded at 1080 × 1920 pixels and about 30 frames per s, placed as in the study. The front camera stood to the participant’s right (median 31°, IQR 21–37°) and was tilted downward (median 20°, IQR 17–22°), as measured from the recordings. The side camera was also on the participant’s right. (C) Characteristics of the analyzed cohort. Blood pressure and heart rate are the mean of each participant’s readings in the analyzed encounters, shown as mean ± SD across participants. Hypertension, BP medication, and home measurement are self-reported. BP ≥130 or ≥80 is the percentage with a mean systolic BP of at least 130 mm Hg or diastolic BP of at least 80 mm Hg. Each value is calculated among the participants with that item recorded (Appendix Table A1). (D) Example Coach-BP verdicts on frames of a volunteer. No study participant is shown. A tick marks a criterion met, a cross a criterion violated, and n/a a criterion not shown for that view. In this preprint, the video frames in panels A and D are shown as line drawings without facial detail. The original images are available from the corresponding authors on request. BP=blood pressure.

## Methods

### Study design and participants

In this prospective, single-center study, we enrolled adults aged 18 years or older attending scheduled appointments at, or employed by, the University of California San Francisco Cardiovascular Prevention Center, San Francisco, CA, USA, between Feb 3 and April 22, 2026. Patients were approached for participation regardless of hypertension status (Appendix) and had to be able to sit upright in a chair. At each encounter, study staff performed a research BP measurement with an Omron 10 Series monitor (BP7465, Omron Healthcare, Kyoto, Japan), a validated upper-arm oscillometric device, before the participant’s appointment. The research reading was not used in care.

Participants completed a four-item intake questionnaire on history of hypertension, antihypertensive medication, home BP monitoring, and previous instruction in measurement technique. Age, sex, gender, race, and ethnicity were not collected, because the protocol was approved as a minimal-risk study that used no protected health information after recruitment. Reporting follows STARD-AI,^17^ with STROBE^18^ for the prevalence analysis and TRIPOD+AI^19^ for model development. The University of California San Francisco institutional review board approved the study (IRB 25-45067), and all participants gave written informed consent.

### Coaching procedure

Each visit began with a baseline reading without instruction on technique. When staff observed incorrect positioning, they gave corrective feedback following the AHA’s home BP measurement guidance^20^ and repeated the measurement in the same sitting. Participants whose technique appeared correct were generally not re-measured. Feedback was delivered by four study-team members (RB, JS, JN, and AJA), two researchers trained in AHA measurement technique by a study cardiologist (JAL-M) and two nurse practitioners. Coach-BP was not used for real-time analysis or feedback during the study. Participants had been seated for at least 5 min before the baseline reading while the study was explained and consent obtained. A repeat reading was taken about 1–2 min later.

### Video acquisition

Each encounter was recorded for about 30–60 s per view with two tablet cameras (Figure 1B), one in front of and one beside the seated patient, both on the patient’s right. Placement was not fixed (Appendix). The front view was used for the arm, legs, feet, and talking criteria and the side view for back supported, which was also tested from the front.

### Landmark extraction

MediaPipe (BlazePose)^13,14^ located 33 body landmarks, such as the shoulders, knees, and ankles, in two and three dimensions in each frame. Up to 30 evenly spaced frames per view were summarized by each landmark’s mean position, its SD, and its mean visibility score. Each landmark’s position in the image was then measured from the center of the patient’s shoulders, in units of the patient’s torso size. The arm, legs, and feet models are therefore not affected by where the patient sits in the frame or how far they are from the camera (Appendix Table A2).

### Criterion definitions and models

Each criterion was scored as met or violated. For each criterion except no talking, we trained an L1-regularized (lasso) logistic regression model with balanced class weights (scikit-learn)^21,22^ on the landmark variables of the relevant view. The feet model also used the output of the legs model. The back model, trained on both views so that it could be tested from the front, also used whole-image features from DINOv2.^15^ Model settings were tuned using the training participants only. Legs and feet models were also trained on left-right mirrored copies of each recording. Scores were converted to verdicts at a threshold of 0·5 fixed before evaluation (Appendix). For no talking, we used TalkNet,^16^ a pretrained audio-visual model that detects whether a visible face is speaking, without fine-tuning. An encounter was classed as talking when TalkNet’s highest score for the patient’s face exceeded zero, its default threshold.

The AHA arm recommendation comprises two conditions, the arm resting on a support and the cuff at heart level. Both panels scored them separately, and the criterion was met only when both were satisfied. Because the cuffed arm varied between encounters, an open-vocabulary object detector (OWLv2)^23^ located the cuff in each front-view video without using clinician ratings. The landmarks were mirrored when the cuff was on the right, so that the cuffed arm was always on the same side.

### Rule-based baselines

To measure what Coach-BP adds, each criterion was also assessed with a rule-based baseline, a decision rule using one or two measurements from the same recording. Back supported was judged by a preset cut-off on the angle between the trunk and the chair backrest, and legs uncrossed by whether the ankles or knees had swapped sides relative to the hips. Our earlier arm and feet rules had been tuned on this cohort, so their baseline was instead the single measurement and cut-off that best separated met from violated in the other participants. For no talking, the baseline was a cut-off on the proportion of the clip with both speech and lip movement (Appendix). Unlike Coach-BP, which gives a score for each encounter, the baselines return only met or violated, so they were compared on balanced accuracy, not AUROC.

### Reference standard

Each encounter was scored from the video by one reviewer from each of two panels of three clinicians. The first panel, a cardiologist and two nurse practitioners from the study team, provided the ratings used in development. The second, two nurse practitioners and a cardiology fellow with no involvement in the study, rated independently. Each reviewer in a panel scored different encounters. Reviewers did not see Coach-BP outputs, second-panel reviewers did not see the first panel’s ratings, and reviewers were not masked to whether an encounter followed coaching.

### Evaluation and statistical analysis

All models were evaluated by leave-one-participant-out cross-validation,^24^ testing each participant with models trained on the others. Performance was summarized by the area under the receiver operating characteristic curve (AUROC) for the scores and by balanced accuracy, the mean of sensitivity and specificity, for the verdicts at the prespecified threshold. Balanced accuracy, which weights violations and met criteria equally, was preferred to accuracy, which rewards never flagging rare violations. Both were also averaged across the five criteria. When no face or person was detected, the encounter was left out only for the criterion affected.

We estimated 95% CIs by bootstrap, resampling participants 2000 times.^25^ Agreement between the two expert panels served as a benchmark for agreement between clinicians. Because Coach-BP was developed on the first panel’s ratings, Coach-BP and the first panel were each compared with the second panel. Agreement was measured as balanced agreement, which is balanced accuracy with one rater as the reference. It was preferred to Cohen’s κ, which depends strongly on how often each rater flags a violation when violations are rare. Cohen’s κ is given in the Appendix. Proportions are reported with Wilson intervals. Tests were two-sided and not adjusted for multiple testing.

In a secondary analysis of participants with more than one reading, the proportion with any error at the first and last readings was compared with the exact McNemar test. Associations between the four self-reported characteristics and any error at the baseline reading were estimated by multivariable logistic regression. For each error that the first expert panel rated as corrected, we report whether Coach-BP classified the corrected reading as meeting the criterion.

### Model interpretability

To interpret each landmark model, closely related variables (Spearman |ρ| of at least 0·7) were grouped into features. In 2000 refits on random halves of participants,^26^ features selected in at least 70% of refits and with an AUROC of at least 0·55 when used alone (single-feature AUROC) are shown. For back supported, whose image features do not correspond to body positions, we instead measured how accuracy changed when parts of the image were replaced with uniform gray (Appendix).

### Robustness to recording conditions

Robustness was tested by altering each participant’s recording and reassessing it with the same cross-validated model, without retraining. We simulated changes in framing (sideways shifts, changes in apparent body size, and rotation), changes of camera viewpoint, low light, reduced resolution, video compression, motion blur, and shortened recordings, with failure prespecified as a fall in AUROC of more than 0·03 or in balanced accuracy of more than 0·05, with the paired 95% interval excluding zero (Appendix).

## Results

### Cohort and prevalence of technique errors

Ten of the 418 recorded encounters were excluded because ratings were missing or the video was unusable (Appendix). The analysis included 408 encounters from 225 of 227 participants. Of the 222 who answered the intake question, 112 (50%) reported a history of hypertension (Figure 1C).

Of the 223 participants whose first reading was analyzed, 138 (62%) had at least one technique error and 54 (24%) two or more (Figure 2). The most common errors were the arm not supported at heart level (76 [34%]) and feet not flat (65 [29%]), followed by lack of back support (42 [19%]), crossed legs (24 [11%]), and talking (11 [5%]). Of 148 participants who routinely measured BP at home, 87 (59%) had at least one error. No self-reported characteristic was associated with having an error after mutual adjustment (Appendix).

**Figure 2:**
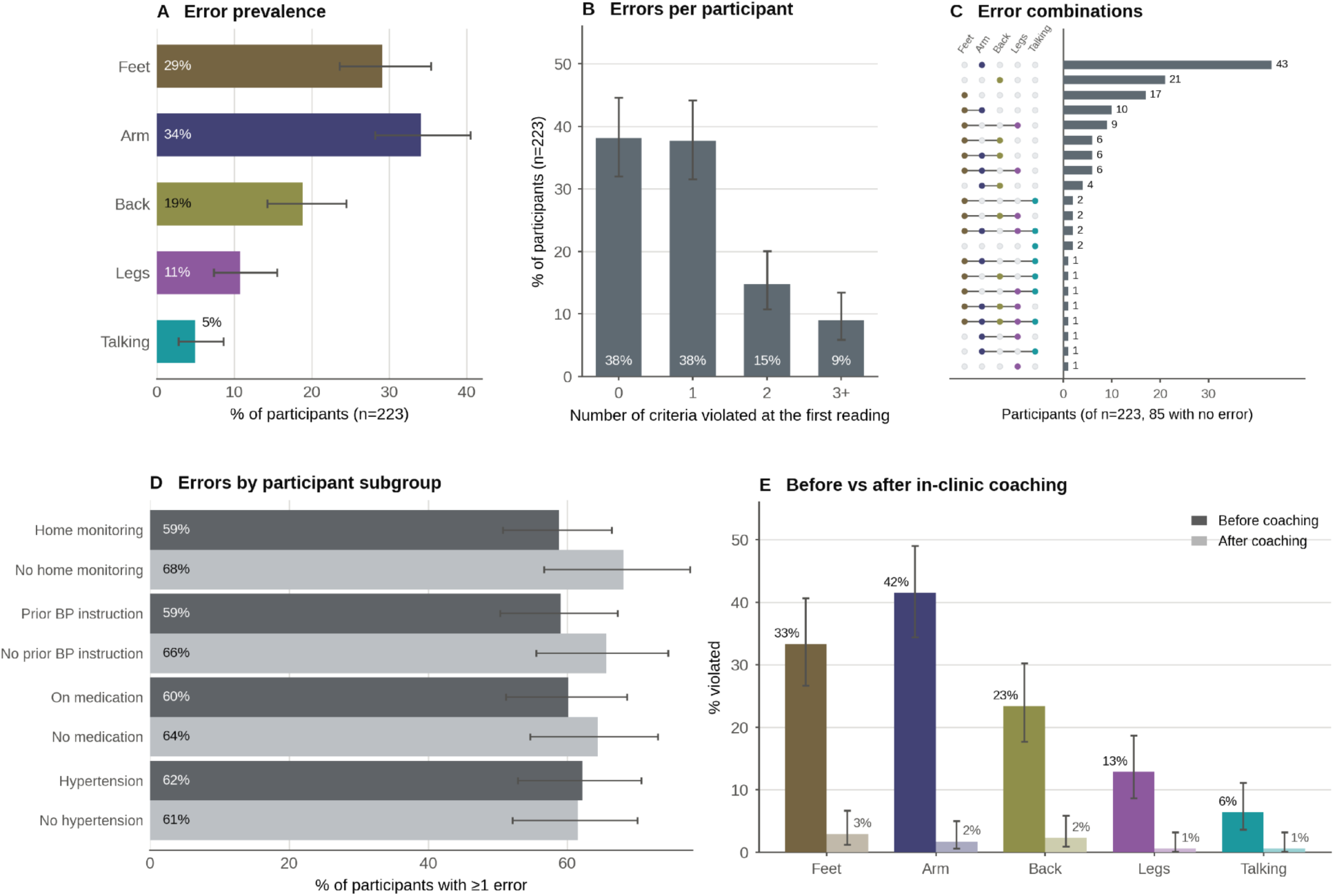
Technique errors at the baseline reading. Data are from the 223 participants whose first reading was included in the analysis (Appendix Figure S1). An arm error means the arm was unsupported or the cuff was not at heart level. (A) Prevalence of each error. (B) Number of criteria violated at each participant’s first reading (three or more in 21 participants). (C) Combinations of errors. (D) Percentage of participants with at least one error, by self-reported characteristic, among those who answered each question (home monitoring 148 yes and 72 no, previous instruction 127 and 93, antihypertensive medication 118 and 101, and history of hypertension 111 and 109). (E) Violation rate for each criterion before and after coaching by staff, at the baseline and final readings of the 171 participants with a repeat reading. Error bars show 95% CIs.

### Detection accuracy

Against the expert ratings used in development (Figure 3A–C; Appendix Table A3), mean AUROC across the five criteria was 0·93 (95% CI 0·90–0·94) and mean balanced accuracy 0·88 (0·85–0·90). Performance was highest for legs uncrossed (AUROC 0·97 [95% CI 0·94–0·99], balanced accuracy 0·92 [0·85–0·98]) and no talking (0·97 [0·93–1·00] and 0·93 [0·85–0·97]), followed by back supported (0·92 [0·85–0·98] and 0·91 [0·86–0·95]), feet flat (0·91 [0·86–0·95] and 0·84 [0·78–0·89]), and the arm criterion (0·86 [0·81–0·91] and 0·78 [0·72–0·84]). Results were similar for baseline readings alone and in encounters on which the two panels agreed (Appendix). Coach-BP gave no talking verdict in seven encounters, where no face was tracked, and no back verdict in two of these, where the participant was not detected. Counting these encounters as errors would lower balanced accuracy by at most 0·011 and AUROC by at most 0·022 (Appendix).

**Figure 3:**
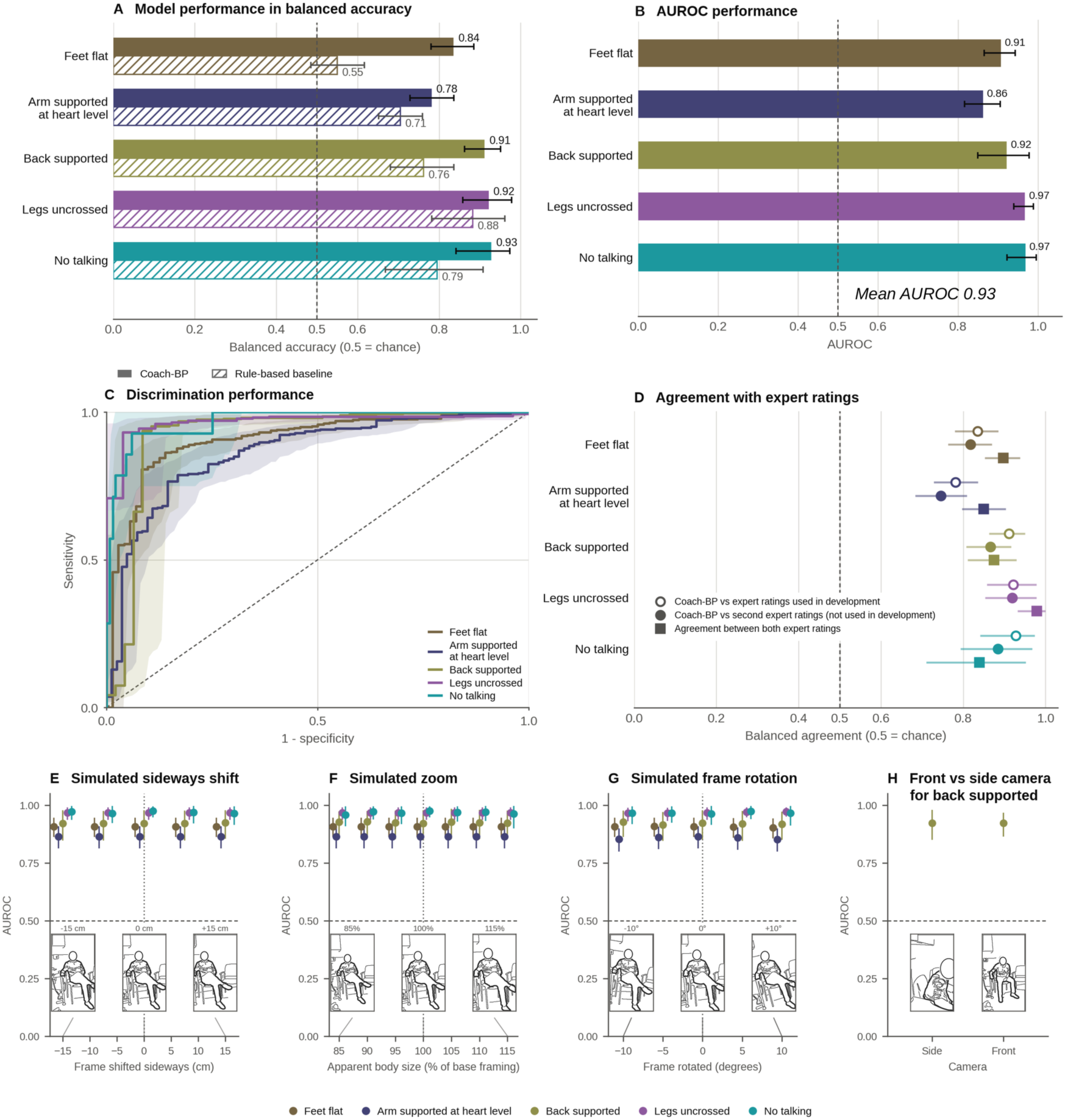
Detection performance and robustness. Coach-BP was tested by leave-one-participant-out cross-validation. In panels A–C and E–H, accuracy is measured against the expert ratings used in development. (A) Balanced accuracy of Coach-BP and of the rule-based baseline for each criterion. (B) AUROC for each criterion, with a mean of 0·93. (C) Receiver operating characteristic curves with pointwise 95% bands. (D) Balanced agreement with expert ratings. The mean was 0·88 for Coach-BP against the ratings used in development. Against the second panel’s ratings, which were not used in development, means were 0·85 for Coach-BP and 0·89 for the first panel. Each encounter was labeled by one rater from each panel. (E–G) Robustness to framing, shown as AUROC when the held-out participant’s recording was shifted sideways (E), zoomed (F), or rotated (G), with the models held fixed. Arm supported, legs uncrossed, and feet flat use positions measured relative to the body, so moving or resizing the image cannot change them. For back supported and no talking, which use the image itself, each change was applied to the frame and compared with an unaltered central crop of 80% of the frame (75% for rotation). Frames show a volunteer who was not a study participant. (H) Camera position, shown as AUROC of the same back supported model from the side and from the front camera, with an example frame of each view. Intervals are 95% CIs. In this preprint, the frames in panels E–H are shown as line drawings without facial detail. The original images are available from the corresponding authors on request. AUROC=area under the receiver operating characteristic curve.

### Comparison with rule-based baselines

Where both gave a verdict, Coach-BP matched or improved on the rule-based baseline for all five criteria and raised mean balanced accuracy from 0·74 to 0·87 (Figure 3A), a difference of 0·13 (95% CI 0·09– 0·18). Gains were clear for feet flat, the arm criterion (0·08 [0·01–0·14]), back supported, and no talking. The difference was 0·03 (−0·11 to 0·17) for legs uncrossed, whose rule already performed well (Appendix).

### Model interpretability and feature importance

Most features corresponded to what a clinician would assess (Figure 4). For the arm criterion, the feature selected in every refit was the height of the cuffed hand above the hip midpoint (f1, single-feature AUROC 0·81 [95% CI 0·74–0·87]), with the hand and elbow positioned forward and out to the side (f3– f5), the pattern of an arm resting on a surface. The second most selected, heel detection confidence (f2), reflects how clearly the lower body was seen. Features for legs uncrossed were the sideways positions of the feet and knees (f6, f7), foot height (f8), and stillness of the near heel (f9, 0·93 [0·89–0·96]), and for feet flat the sideways and forward positions and height of the feet (f10, f11, f13, f14) and the legs-model output (f12).

**Figure 4:**
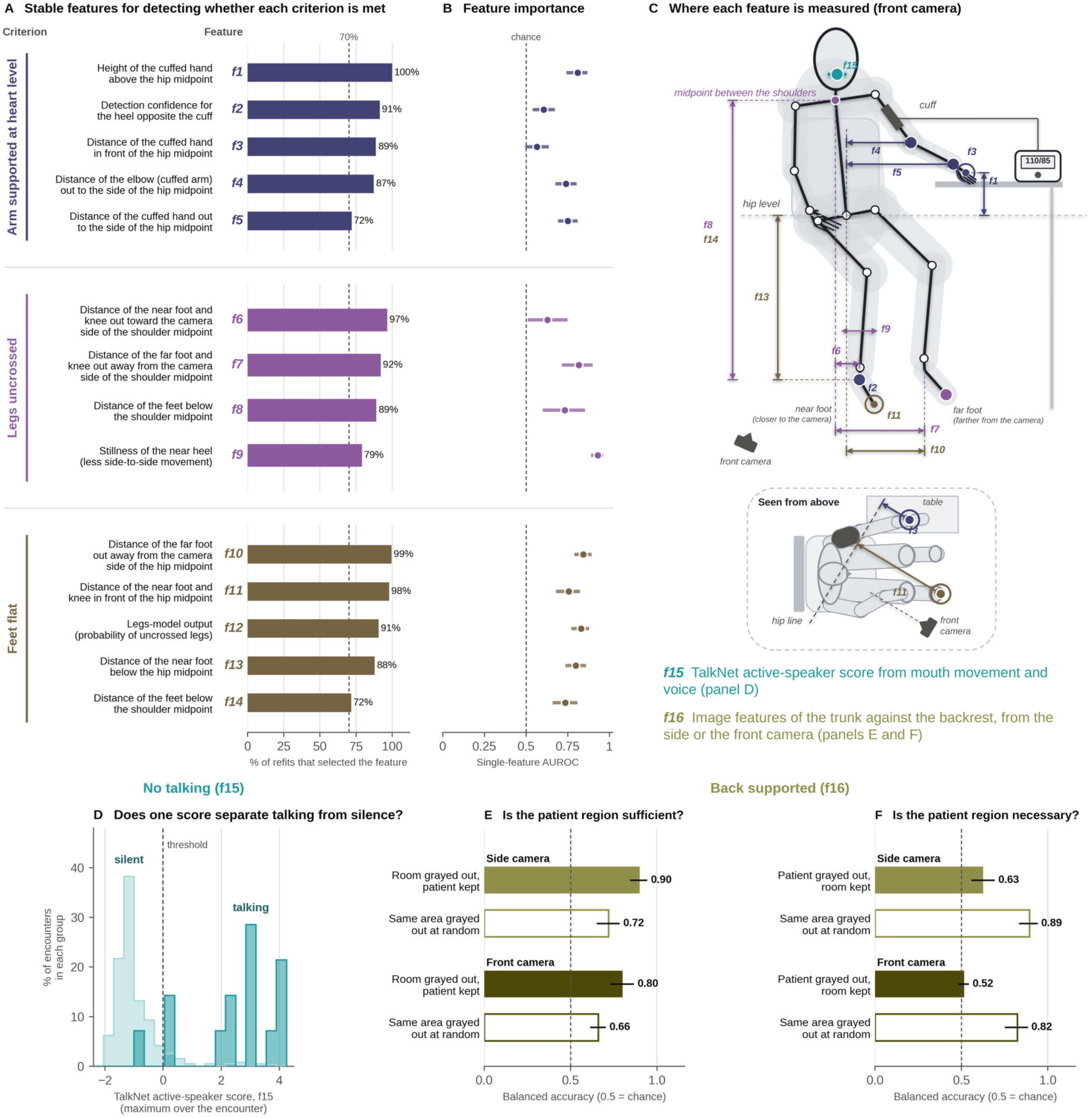
Model interpretability and feature importance. (A, B) One row per feature, grouped by criterion (arm supported at heart level, legs uncrossed, and feet flat). Features f1–f14 are numbered by selection frequency within each criterion and drawn in panel C. Each feature groups landmark variables that measure the same thing, with every pair correlating at least 0·7. For example, f1, the height of the cuffed hand, combines eight hand landmark variables. The grouping is for interpretation only, and the models use each landmark variable separately. Each feature is named in the direction that goes with the criterion being met. (A) Percentage of 2000 refits on random halves of participants in which the feature was selected (dashed line, 70%). (B) Single-feature AUROC, the AUROC of the feature used alone (dashed line, chance), with 95% CIs. (C) Where each feature is measured, drawn from the median pose of the encounters that met every criterion, as seen by the front camera. Each mark sits on the feature’s most frequently selected landmark, and ringed points (f3, f11) measure depth and are shown in the view from above. Near and far refer to the front camera (drawn in C), which stood on the patient’s right in 388 (95%) of 408 encounters. Arm features are named by the cuffed side, detected from the video. (D) Maximum TalkNet speaking score over the encounter (f15) in the 14 talking and 387 silent encounters, with the prespecified threshold of zero. (E, F) Back supported relies on image features that do not correspond to body positions (f16), so parts of the image from each camera were grayed out and the features recomputed. In E everything outside a box enclosing the patient and backrest was grayed out, and in F the patient box was grayed out, each compared with graying out the same area at a random position. AUROC=area under the receiver operating characteristic curve.

The arm criterion’s two conditions diverged in 63 (15%) of 408 encounters, and cuff height was harder to classify than arm support (Appendix).

For back supported, graying out everything except the patient and backrest lowered balanced accuracy from 0·94 to 0·90 (95% CI 0·84–0·94), whereas graying out the patient reduced it to 0·63 (0·56–0·69), and the front view showed the same pattern (Figure 4E, F). No talking rests on one number, the maximum TalkNet score, which classified 13 of 14 talking and 359 of 387 silent encounters correctly (Figure 4D).

### Agreement with independent raters

Mean balanced agreement of Coach-BP with the second expert panel was 0·85 (95% CI 0·82–0·88), compared with 0·88 (0·85–0·90) with the first panel and 0·89 (0·85–0·92) between the two panels (Figure 3D). Coach-BP’s agreement with the second panel was slightly lower than that between the panels (difference −0·041 [95% CI −0·074 to −0·004]), and lower for the arm criterion (−0·10 [−0·18 to −0·03]) and feet flat (−0·08 [−0·14 to −0·02]).

### Robustness to recording conditions

Sideways shifts and changes in apparent size left the landmark models unchanged, and no change in framing altered any model’s AUROC by more than 0·02 (Figure 3E–G). Under simulated changes of camera viewpoint, AUROC for legs uncrossed and feet flat stayed within 0·82–0·99, whereas for the arm criterion it fell from 0·84–0·88 between head-on and 60° to the patient’s right to 0·73–0·81 with the camera on the patient’s left (Appendix Figure S3). The same back model was similarly accurate from the front camera, placed about 30° to the patient’s right, as from the side camera (AUROC 0·92 and 0·92, balanced accuracy 0·88 and 0·91, Figure 3H, Appendix Table A6), but less robust to frame rotation (Appendix). A back model trained on one view alone was much less accurate from the other camera (Appendix Table A6). No model failed under reduced resolution or motion blur. Severe low light degraded the trained models, severe compression degraded the legs and back models, and quarter-length recordings degraded the legs model (Appendix).

### Technique before and after coaching

The 171 participants with a repeat reading more often had a baseline error than the 52 without one (75% vs 19%). Among them, the number with any error fell from 128 (75%) to 12 (7%), a difference of −67·8 percentage points (95% CI −74·8 to −60·8, p<0·0001). All 43 participants correct at baseline remained correct. By the second panel’s ratings, the number fell from 114 (67%) to 16 (9%) of the 170 rated at both readings. When an error was corrected, Coach-BP classified that reading as meeting the criterion in 172 (91%) of 189 cases (Appendix). At the repeat reading, Coach-BP flagged at least one error in 40 (24%) of the 170 with all five verdicts, compared with 12 (7%) by the first panel.

## Discussion

Coach-BP detected errors in all five AHA criteria from ordinary clinic video, with a mean AUROC of 0·93, ranging from 0·86 for the arm criterion to 0·97 for legs uncrossed and no talking. Such errors were common, affecting 62% of participants at baseline, including 59% of those who routinely measured BP at home. These estimates describe patients positioning themselves without instruction while waiting for their appointment, which is closest to unsupervised home measurement, rather than routine clinic measurement, in which staff are expected to position the patient. Back support, for which we had placed a second camera, was assessed about as well from the front camera, so a single front camera, placed as in the study, could assess all five criteria, although it was more sensitive to frame rotation.

Our prevalence estimates are consistent with studies that assessed technique by observation or video. Among 100 patients with hypertension filmed while measuring their own BP in primary care, only 3% made no error, judged against a broader checklist than ours.^27^ Such assessment depends on trained observers. The only previous automated check of seated posture we found, a sensor-equipped chair,^28–30^ requires dedicated equipment and does not assess the arm or feet. We are not aware of a previous system that assesses all five criteria from ordinary video and has been evaluated against clinician review. Because it returns a verdict for each criterion, Coach-BP is designed to show what needs correcting.

The arm criterion was the least accurately classified, and Coach-BP’s agreement with the second panel was lowest for it (balanced agreement 0·75, compared with 0·85 between the two panels). With the cuff on the right arm, the camera usually stood on that side, and there the arm criterion was less accurate (AUROC 0·76 vs 0·90) and clinicians also agreed less (0·77 vs 0·88) (Appendix). Adding an estimate of heart level, depth information, or the side camera did not improve the arm criterion, and for cuff height alone the best model agreed with the second panel as well as the two panels agreed with each other (balanced agreement 0·71 for both, Appendix). This suggests that cuff height is hard to judge from video, for clinicians and models alike. A more precise definition of heart level, scored separately from arm support, would help both.

Coach-BP assesses the conditions of a reading, not the BP value. A missed error lets through a reading that is usually biased upwards, whereas a false alert costs an unnecessary repeat reading but, if frequent, could erode trust. Here, 123 (31%) of the 401 encounters with all five verdicts had at least one false alert, and 28 of 41 talking alerts were false, so thresholds for coaching remain to be set. Withholding verdicts with a score between 0·3 and 0·7 raised mean balanced accuracy from 0·88 to 0·93, but 125 (31%) of 401 encounters then received all five verdicts (Appendix Table A8). Deployment should show each verdict with the image it is based on, keep a human check in clinic use, and audit verdicts against human review. Video also captures faces, speech, and, at home, other household members. Because the body-tracking model was designed for mobile devices,^14^ a deployed system could analyze video on the device and store only the verdicts.

The substantial reduction in technique errors following brief staff-delivered coaching suggests that most errors are readily correctable. Future studies should evaluate whether Coach-BP can reproduce these improvements through real-time automated feedback in both clinic and home settings, including determining the type, intensity, and frequency of coaching needed to achieve and sustain proper measurement technique. More broadly, establishing automated technique verification as a standard component of home BP monitoring will require validation across diverse home environments, camera types, lighting conditions, furniture, clothing, and patient populations. Such studies should evaluate detection accuracy and the feasibility and acceptability of integrating automated coaching into routine home BP monitoring.

### Limitations

Coach-BP was developed and evaluated in one cohort by cross-validation, at a single center, with the same tablet model for every recording. Simulated changes in framing, camera viewpoint, and recording quality partly address this, but validation across sites, devices, and homes is needed. No demographic or anthropometric data were collected, so performance by age, sex, race and ethnicity, skin tone, or body size remains to be examined. Pose and face detection models can perform differently across these groups, and future studies should record these characteristics. Talking and crossed legs were uncommon (14 and 26 encounters), so their confidence intervals are wider, and larger samples would narrow them. The coaching comparison was observational, in participants whom staff re-measured because they saw an error. The second panel, which had no role in coaching, found a similar fall in errors. Thus, automated coaching is expected to achieve the same, which remains to be established.

## Contributors

RB, JAL-M, and RA-A designed the study, developed Coach-BP, performed the analysis, and wrote the first draft of the manuscript. RB collected data with contributions from JN, AJA, JS, JAL-M, and RA-A. MJP contributed to interpretation of the findings. All authors reviewed and revised the manuscript.

## Data Availability

The video recordings show participants' faces and cannot be shared under the terms of participant consent. De-identified data that support the findings of this study may be made available on reasonable request to the corresponding author for non-commercial research, subject to approval by the University of California, San Francisco and a data use agreement. The study protocol and the consent form are available on request.

## Acknowledgments

We thank the patients and staff of the University of California, San Francisco Cardiovascular Prevention Center who contributed to this study. In particular, we thank Sara Mendoza and Ava Goei-Vujovich for their tremendous contributions to participant recruitment and study coordination, and the medical assistants of the center for arranging rooms and supporting the study in clinic. We thank Emily Clarke, Kimberly Trinh, and Simon Feseha for independently rating the recordings. We also thank Kashvi Gupta for help with recruitment and recording. During this work the authors used Claude (Anthropic) as an aid in writing analysis code, searching the literature, and editing the manuscript text. The authors directed this use, reviewed and verified all code, results, references, and text, and take full responsibility for the content of the publication.

## Code availability

The analysis code will be made available to academic researchers for non-commercial use.

## Appendix

### 1. Participants, exclusions and coaching

In this prospective, single-center study, we enrolled adults aged 18 years or older attending scheduled appointments at, or employed by, the University of California San Francisco Cardiovascular Prevention Center, San Francisco, CA, USA, between Feb 3 and April 22, 2026, over 30 clinic days. Eligible participants were able to sit upright in a chair for BP measurement and to give written informed consent. People who declined video recording were not enrolled. Patients were approached as a convenience sample, in practice most often when they had arrived early and faced a substantial wait before their appointment, regardless of whether they had a diagnosis of hypertension. Twelve participants were recorded as clinic staff. Patients who declined were not recorded, so the number approached is unknown. No formal sample size calculation was made, and the number of violations available for each criterion is reported with each estimate. At each encounter, study staff performed a BP measurement for research purposes with an Omron 10 Series upper-arm oscillometric monitor (BP7465, Omron Healthcare) and its supplied cuff for arm circumferences of 22–42 cm. Measurements were taken in a room at the center during the waiting period before the participant’s appointment. It did not replace or alter the participant’s clinical measurement, and study readings were not entered in the medical record or used in care. Video was recorded for the study, reference labels were assigned afterwards from the recordings, and Coach-BP was developed and internally validated in the same cohort.

Ten of the 418 recorded encounters were excluded before analysis (Table A1, Figure S1). Four were not rated by the first expert panel, in four the clip shown to the reviewers did not show the participant (Section 2), one had no front recording, and in one another person stood between the front camera and the participant. The exclusions left two participants with no analyzed encounter, so the analysis cohort comprised 225 participants and 408 encounters. Coach-BP returned a verdict for every analyzed encounter for feet flat, the arm criterion, and legs uncrossed. For no talking it returned none in seven encounters, in which no face was tracked. In two of these seven it also returned none for back supported from the side recording, because the pose model did not detect the participant in it. These encounters were omitted for that criterion only, leaving 401 encounters for no talking and 406 for back supported from the side recording. One of the two without a verdict from the side recording had a violation in the expert ratings, and none of the seven without a verdict for no talking did. Counting these missing verdicts as errors would lower balanced accuracy by 0·011 for back supported from the side recording and by 0·008 for no talking, and in the worst case would lower AUROC by 0·022 and 0·017. This study is reported in accordance with the STARD-AI reporting guideline for diagnostic accuracy studies using artificial intelligence.^1^ The descriptive analysis of error prevalence additionally follows STROBE,^2^ and reporting of model development and internal validation follows TRIPOD+AI.^3^ The study was approved by the University of California San Francisco institutional review board (IRB 25-45067), and all participants provided written informed consent. No adverse events occurred. The cohort was a convenience sample at one prevention clinic, and 185 of the 408 analyzed encounters were readings repeated within the visit, generally after coaching, so the frequency of violations may differ from that in home use.

**Appendix Figure S1:**
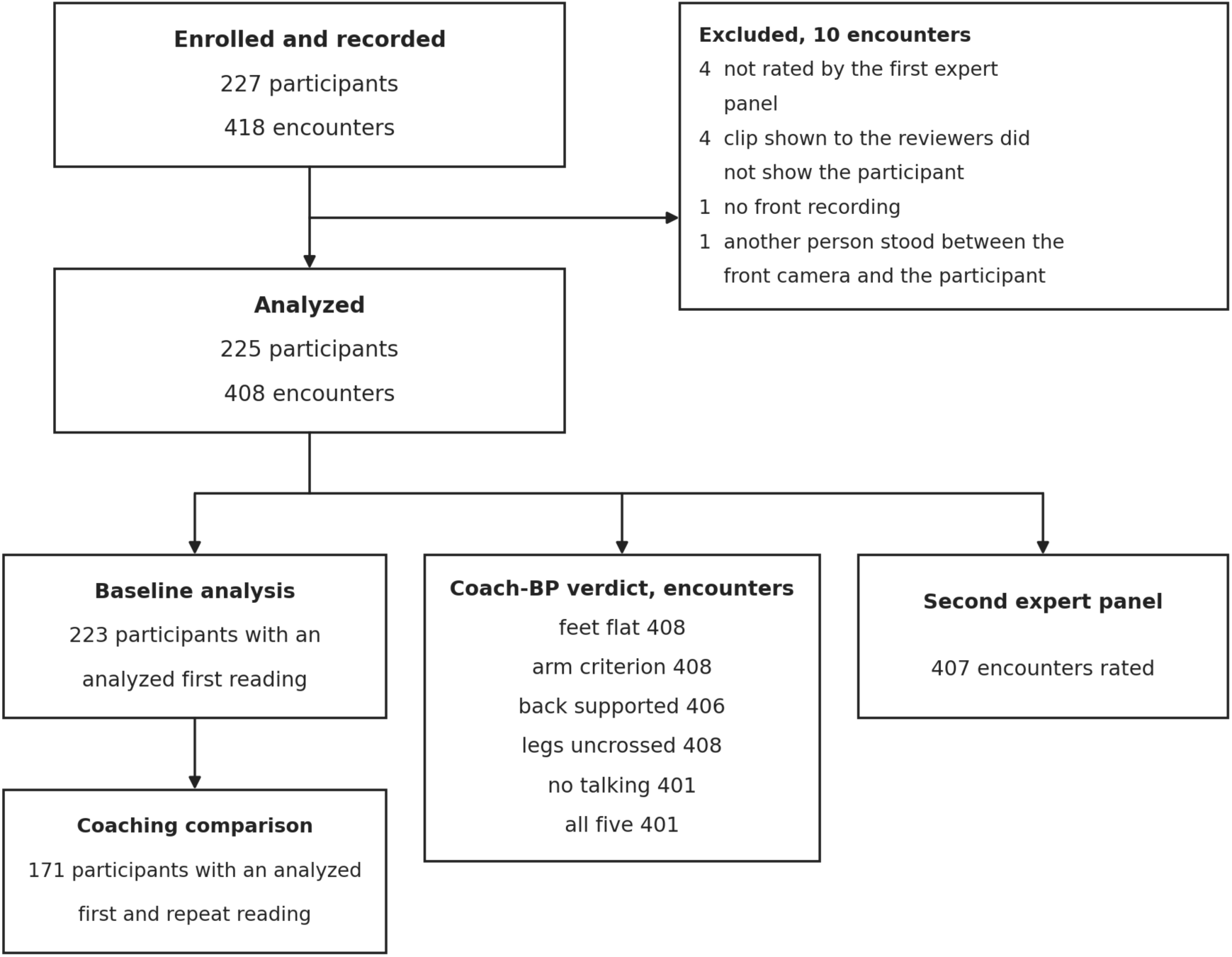
Flow of participants and encounters. Of the 227 participants enrolled and 418 encounters recorded, 10 encounters were excluded, which removed two participants entirely and left 225 participants and 408 encounters for analysis. The lower boxes give the numbers in each analysis (Table A1). Two of the 225 analyzed participants had no analyzed first reading and are not in the baseline analysis.

**Table A1:** Participants and encounters in each analysis.

| Analysis | Number | Reason for any difference |
| --- | --- | --- |
| **Participants** |  |  |
| Enrolled | 227 |  |
| Analyzed | 225 | The excluded encounters left two participants with no analyzed encounter |
| Answered the intake item on history of hypertension | 222 | Three did not answer. Home monitoring and previous instruction were also answered by 222, and antihypertensive medication by 221 |
| Blood pressure and heart rate recorded (Figure 1C) | 213 and 205 | No reading was available for 12 participants, and eight others had no heart rate |
| First reading analyzed (baseline analysis) | 223 | For two participants the first reading was excluded (one not rated by the first expert panel, one with no front recording) and a later reading was analyzed |
| First reading analyzed and all four intake items answered (adjusted analysis) | 219 | Four of the 223 left at least one item unanswered |
| First and repeat reading analyzed (coaching comparison) | 171 | The other 52 of the 223 had no analyzed repeat reading. The last reading was used where there were several |
| Coaching comparison with the second expert panel, and repeat reading with all five Coach-BP verdicts | 170 each | For one of the 171, the second panel did not rate one recording, and for another one verdict was missing at the repeat reading |
| **Encounters** |  |  |
| Recorded | 418 |  |
| Excluded | 10 | Not rated by the first expert panel (four). Clip shown to the reviewers did not show the participant (four). No front recording (one). Another person stood between the front camera and the participant (one) |
| Analyzed | 408 |  |
| With a Coach-BP verdict | Feet flat 408, arm criterion 408, back supported 406, legs uncrossed 408, no talking 401 | Missing verdicts are explained in the text above |
| With all five Coach-BP verdicts | 401 | Seven encounters lacked at least one verdict |
| Rated by the second expert panel | 407 | The front video of one encounter did not load for the second-panel reviewer |
| With a Coach-BP verdict and a second-panel rating (Table A4) | Feet flat 407, arm criterion 407, back supported 405, legs uncrossed 407, no talking 400 |  |
| On which the two expert panels agreed | Feet flat 384, arm criterion 363, back supported 387, legs uncrossed 405, no talking 391 | Among the encounters in the row above. For back supported from the front recording, 389 of the 407 |
| First readings with a Coach-BP verdict (baseline readings alone) | Feet flat 223, arm criterion 223, back supported 221, legs uncrossed 223, no talking 218 | Among the 223 analyzed first readings |
| Rule-based baseline, with a verdict (and paired with Coach-BP) | Feet flat 408, arm criterion 408, back supported 406, legs uncrossed 368, no talking 363 (361 paired) | The rule returned no output for the others |
| Cuffed arm: detected, staff record, both (comparison) | 404, 396, 392 | The detector did not find the cuff in four of the 408, and staff recorded no arm for 12 |
| Graying-out analysis for back supported (Figure 4E, F) | 288 from the side camera, 390 from the front camera | The pose model had to locate the patient in every sampled frame of that view so that the boxes could be placed |
| Framing analysis for no talking (Figure 3E–G) | 401 | Encounters with a tracked face |
| Degraded recordings (Table A5) | Arm criterion 408, legs uncrossed 408, feet flat 408, back supported 406 | Encounters with a Coach-BP verdict for that criterion |
| Camera tilt (Figure S3C) | 388 | Recordings with at least ten vertical edges from which to estimate tilt |
| Median pose (Figure 4C) | 252 | Encounters that met every criterion |
| Back supported from the front recording (Section 11) | 408 | Every analyzed encounter had a front recording |

Each analysis used every participant or encounter with the data it needed, so the numbers differ between analyses. Back supported is one model tested from each camera, and its numbers are for the test from the side camera unless the row says otherwise.

Each visit began with a naturalistic baseline reading, during which no instruction on measurement technique was given. Where the member of staff performing the measurement observed incorrect positioning, they provided corrective feedback and repeated the measurement within the same sitting. Participants whose technique appeared correct at the baseline reading were generally not re-measured. Feedback followed the positioning instructions of the American Heart Association’s Home Blood Pressure Measurement Instructions,^4^ which direct the patient to sit upright with the back supported, feet on the floor and legs uncrossed, to rest the arm comfortably on a flat surface at heart level, and to stay relaxed and not talk. Staff directed the feedback at whichever element was incorrect. The time seated before the baseline reading was not measured, because the study assessed measurement technique and BP values were not an outcome. Participants had been seated in the waiting room, and were then seated for at least 5 min while the study was explained and written consent was obtained. The chair had a backrest and a surface on each side for the arm, a desk on the participant’s right and a side table on the left, of similar and fixed height. The repeat reading was taken about 1–2 min after the baseline reading. Feedback was delivered on most study days by one researcher (RB) and on the remaining days by a second researcher (JS) or one of two cardiology nurse practitioners (JN and AJA). The two researchers had been trained in AHA measurement technique by a study cardiologist (JAL-M). Two people who are not authors, a nephrology fellow and a clinic medical assistant, each helped with recruitment and recording on one or two study days. Corrective feedback and repeat measurement were study procedures rather than routine clinical care. Coaching was delivered by study personnel and not by Coach-BP. The system was not used during data collection, and its outputs were not available to staff or to the clinician reviewers.

### 2. Video acquisition and camera geometry

Each BP encounter was recorded with two tablet cameras (iPad Air 11-inch [M3], Apple, Cupertino, CA, USA, running iPadOS 26.3, 1080 × 1920 pixels, about 30 frames per s), one in front of and one beside the seated patient, both on the patient’s right, for about 30–60 s per view. Placement was not fixed. Measured retrospectively from the recordings, the front camera viewed the patient from a median of 31° to their right (IQR 21–37°) and pointed a median of 20° downward (IQR 17–22°). The front-view video was used to assess arm supported at heart level, legs uncrossed, feet flat on the floor and no talking. Back supported was assessed from the side-view video and, separately, from the front-view video (Section 11). Coach-BP was applied offline to the clips described below, and its outputs were not shown to participants or staff. Real-time use was not evaluated, nor was analysis on the recording device. For analysis, most recordings were cut to their central 20 s, and the rest were used whole or cut by hand. Of the 816 clips analyzed, two views of each of the 408 encounters, 694 (85%) lasted about 20 s (19–21 s), 29 (4%) were shorter, and 93 (11%) were longer. The reviewers rated these clips, and Coach-BP was run on the same clips. In four encounters the central 20 s did not show the participant, and these encounters were excluded (Section 1). Coach-BP is intended for patients measuring their own blood pressure and for clinic staff taking a measurement, and it requires no clinical or technical expertise beyond placing a camera.

Camera tilt was estimated from the vanishing point of vertical edges in the room (door frames, wall corners, furniture), in three frames per recording, with the focal length estimated from the same images. The estimate changed little with an error of ±20% in focal length or when only the patient’s body was masked. The front camera’s angle to the patient was estimated as the angle of the three-dimensional shoulder line to the camera, and so reflects both where the camera stood and how the patient sat. Both cameras were on the patient’s right whichever arm was cuffed (front camera in 97% of left-arm and 91% of right-arm sessions, side camera in 95% and 94%), but the angle to the front camera was larger with the cuff on the left arm (median 33°, IQR 28–38°) than on the right (16°, 7–30°), consistent with patients turning slightly toward the cuffed arm. These values describe how placement varied rather than a protocol.

### 3. Landmark variables

We extracted body landmarks with MediaPipe (BlazePose),^5,6^ an open-source on-device pose-estimation framework that returns 33 anatomical landmarks in each video frame, each with a visibility score. Landmark positions were represented both in two-dimensional image coordinates and in three-dimensional coordinates,^7^ in approximate meters, centered on the hip midpoint, extracted from the same frames. Up to 30 frames per view were sampled at evenly spaced intervals across each recording. For each encounter, landmark coordinates were summarized across frames as the mean and SD of each coordinate and the mean visibility score, giving 165 two-dimensional landmark variables (33 landmarks × five summary statistics) and 198 three-dimensional landmark variables (33 landmarks × six). Two-dimensional coordinates of both views were then expressed relative to the patient’s body rather than the video frame. Each coordinate was measured from the midpoint of the two shoulders and divided by the distance from one shoulder to the opposite hip, and within-recording SDs were divided by the same distance. This makes the arm, legs and feet models exactly invariant to where the patient sits in the frame and to the patient’s distance from the camera, but not to rotation of the frame or a change of viewpoint, which were tested by simulation. The inputs of the back model also include scene embeddings of the whole image, which are not referenced to the body, so its robustness to framing was tested on real image crops (Section 9).

The shoulder-to-opposite-hip distance was chosen as the scale because it varies less between recordings than shoulder width (coefficient of variation 0·140 vs 0·214), which rotation of the body foreshortens.

### 4. Models

Each of the five AHA criteria was scored as a binary outcome (criterion met vs violated). We evaluated rule-based baselines, supervised models and, for back supported and no talking, feature-specific models. Reviewers also recorded whether the back was straight, whether the patient remained still and whether the cuff was on bare skin. These items were outside the five criteria that the study set out to assess and are not analyzed, except back straight in Section 11.

#### Rule-based baselines

Back supported was classified by the angle between the trunk and the chair backrest identified in the side-view video, and legs uncrossed by the relative ordering of the left and right knees and ankles, with thresholds fixed a priori from ergonomic considerations. For the arm and feet criteria, whose geometric rules had been tuned on the study cohort, the baseline reported is instead the single front-view variable that best separated the classes within each training fold, with its cut-off searched over every candidate value in that fold, evaluated on the held-out participant. For no talking, the baseline was an audio-visual heuristic, the proportion of the clip in which detected speech coincided with movement of the patient’s lips. Its threshold had been selected on the study data, and it was run on the front clip that the reviewers saw with that threshold unchanged.

#### Supervised machine-learning models

For each criterion we trained an L1-regularized logistic-regression classifier (LogisticRegressionCV with the liblinear solver and balanced class weights)^8,9^ on the encounter-level landmark variables. The legs model used front-view two-dimensional variables, and the feet model used front-view two-dimensional and three-dimensional variables and the output of the legs model. The arm model used front-view variables aligned to the cuffed side (described below). The inputs of the back model were the two-dimensional and three-dimensional variables of a recording and its scene embedding, and it was trained on the side and front recordings of every training encounter together, so that one model could be tested from either camera (Section 11). The regularization strength was selected within each training fold, using the training participants only, by five-fold inner cross-validation, stratified by outcome and grouped by participant, over a logarithmic grid of eight values between 10⁻² and 10¹, with AUROC as the selection metric. The inner cross-validation was repeated ten times with different random fold assignments, and out-of-fold predictions were averaged across the repetitions. Random seeds were fixed for every fold assignment and solver, so all results reproduce exactly. Models for legs uncrossed and feet flat were trained with left-right mirrored copies of each training recording, so that they tolerate a camera placed on the patient’s other side. Further augmentation was prespecified only for a criterion that failed a robustness test, and none was adopted (Table A2). Each model produced a score between 0 and 1, converted to a verdict at a threshold of 0·5 fixed before evaluation. With balanced class weights, this threshold treats sensitivity and specificity as equally important. Because of this class weighting, the scores rank encounters but are not calibrated probabilities (calibration is reported below). Choices between feature representations were made by comparing cross-validated performance, so the estimates may be modestly optimistic. The main choices were repeated within the training folds (below). For deployment, each model was refitted on all participants.

#### Feature-specific models

For back supported, pose features were augmented with 768-dimensional embeddings from the pretrained DINOv2 ViT-B/14 model, averaged across uniformly sampled frames of the recording.^10^ For no talking, we used TalkNet, a pretrained audio-visual active-speaker detection model,^11^ without fine-tuning. The patient’s face was detected and tracked with the TalkNet reference pipeline, the largest face track was taken as the patient’s, and frame-level active-speaker scores were summarized as their maximum over the encounter. An encounter was classified as talking when this exceeded zero, TalkNet’s native decision boundary. The maximum was chosen from three summaries after an initial run on 34 encounters that included all 14 then labeled as talking. Any talking during the reading violates the criterion, so the maximum over the reading is the natural summary. In the 401 analyzed encounters with a tracked face, 14 of them with talking, the maximum had an AUROC of 0·97 and, at the cutoff of zero, flagged 13 of the 14 with 28 false alerts among the 387 silent encounters. The 90th percentile had an AUROC of 0·92 and flagged eight of the 14 with four false alerts, and the mean had an AUROC of 0·79 and flagged no encounter.

#### The arm criterion

The AHA recommendation that the measurement arm be supported at heart level comprises two conditions, that the arm rests on a supporting surface and that the cuff sits at heart level. Both rating panels scored these as separate items, and we defined the arm criterion as met only when both were satisfied, consistent with the guideline. The relevant arm is the one wearing the cuff, which varied between encounters. We therefore located the cuff automatically in each front-view video with an open-vocabulary object detector (OWLv2),^12^ applied to a crop of each arm from shoulder to wrist in three frames, and took the arm with the higher cuff score as the cuffed arm. The clinician ratings were not used. In four encounters the detector did not find the cuff, and the left arm, the more common side, was assumed. This matched the staff record in all four. The detected side agreed with the staff record in 354 (90%) of the 392 encounters that had both a cuff detection and a staff record (Table A1). Pose landmarks of encounters with the cuff on the right arm were mirrored, so that the cuffed arm always occupied the same variables, and a single L1-regularized logistic-regression model was trained on the composite label using the mirrored two-dimensional and three-dimensional front-view variables.

The arm criterion combines two conditions that often diverged. In the expert ratings used in development, the arm was supported but the cuff not at heart level, or the reverse, in 63 (15%) of 408 encounters. A small set of hand-built geometric features predicted whether the arm was supported (balanced accuracy 0·69) but was no better than chance for whether the cuff was at heart level (0·50). Trained on cuff height alone, the inputs of the final arm model reached an AUROC of 0·69 (0·60–0·78). Adding a heart-level reference from the shoulder and hip midpoints, landmark depth, or side-camera landmarks did not improve this or the arm criterion. Agreement between the two expert panels on cuff height alone was 0·71 (0·63–0·79), and the best model reached 0·71 with the second panel.

#### Choices repeated within the training folds

Choices that had been made by comparing candidates in this cohort were repeated inside every training fold. In each fit, the candidate with the highest mean AUROC in the inner cross-validation scored the held-out participant, so that the estimate includes the choice. For feet flat, with the choice of whether to give the model the output of the legs model repeated in this way, AUROC and balanced accuracy were unchanged (differences 0·000 [95% CI −0·009 to 0·008] and 0·005 [−0·019 to 0·032]). For back supported, a candidate with both the pose variables and the scene embedding was chosen in 2246 of the 2250 fits for the side camera and in 2245 for the front camera, so that choice was reproduced. The one model for both views was not chosen for the highest cross-validated score. The model trained on front recordings alone had a higher AUROC from the front camera (Table A6), and the one model was adopted because one model served both cameras at the common threshold, under a rule written before its results were seen. The choice by AUROC within folds took a model trained on one view in 1510 (67%) of the 2250 fits for the side camera and in 1919 (85%) for the front camera. Balanced accuracy at 0·5 was then 0·90 from the side camera and 0·84 from the front camera, against 0·91 and 0·88 for the one model (differences −0·009 [−0·043 to 0·016] and −0·033 [−0·089 to 0·022]), with AUROCs of 0·92 and 0·93. For the arm criterion, repeating the choice among 31 input configurations within the training folds did not lower the estimate (balanced accuracy 0·79 against 0·78, difference 0·006 [95% CI −0·043 to 0·054]). Other configurations scored similarly, so the estimate does not depend on this choice.

**Table A2:** Reference frame, invariance and training augmentation for each type of model input.

| Model input (criteria that use it) | Reference and what it is invariant to | Training augmentation |
| --- | --- | --- |
| Front-view 2-D landmark positions (arm, legs, feet) | From the shoulder midpoint, divided by the shoulder-to-opposite-hip distance, so invariant to position in the frame and distance from the camera | Legs and feet with left-right mirrored copies. Arm aligned to the cuffed side |
| Front-view 2-D within-recording SDs (arm, legs, feet) | Divided by the same distance, so invariant to distance from the camera | As above |
| Front-view 3-D landmark positions and SDs (arm, feet) | From the hip midpoint, in approximate meters, so invariant to position and distance | Feet with mirrored copies. Arm aligned to the cuffed side |
| Landmark detection confidence (arm, legs, feet) | The pose model's own score from 0 to 1, which depends on image quality and occlusion, not on position or distance | Legs and feet with mirrored copies, left and right swapped. Arm aligned to the cuffed side |
| Legs-model output (feet) | Probability of uncrossed legs, invariant to position and distance because its inputs are | Legs model with mirrored copies |
| 2-D and 3-D landmark variables of the side or front view (back) | As the 2-D and 3-D front-view positions above, for these inputs only | None. Trained on side and front recordings together |
| DINOv2 scene embeddings (back) | The whole image. Not invariant by construction, but pretraining on cropped and rescaled images makes them tolerant of moderate reframing | None. Trained on side and front recordings together |
| TalkNet active-speaker score (no talking) | A crop that follows the tracked face, so largely invariant to position and size | None. Used without fine-tuning |

Augmentation results are from development runs. Mirrored training copies kept balanced accuracy on the recordings as made and restored it on mirrored recordings (legs uncrossed 0·82 to 0·93, feet flat 0·50 to 0·84). Simulated viewpoint changes and a viewpoint-invariant pose frame for the arm criterion, and darkened copies for back supported, did not meet the criteria for adoption and were not used. 2-D=two-dimensional. 3-D=three-dimensional.

#### Calibration

Because of balanced class weighting, mean scores of the four trained models (0·63–0·82) were lower than the observed proportions meeting the criterion (0·80–0·94). Calibration intercepts were 1·23–2·19 and slopes 0·86–1·38. These values take back supported from the side camera. No talking is excluded because its score is a logit.

### 5. Reference standard

Each encounter was independently scored for the five criteria by one reviewer from each of two three-clinician panels, working from the recorded video rather than by observation at the time of measurement. Review of the recording was chosen so that every reviewer and Coach-BP judged the same clip, and so that a second panel could rate it independently. The first expert panel comprised three clinician reviewers who were members of the study team, and the second expert panel comprised three clinician reviewers who had no involvement in the study. First-panel reviewers were a cardiologist specializing in prevention (JAL-M) and two cardiology nurse practitioners (JN and AJA). Second-panel reviewers were two nurse practitioners in adult cardiology and a clinical fellow in cardiology at the same institution, none of whom had any other part in the study. Within each panel, reviewers scored disjoint batches of encounters, so each encounter received one first-panel rating and one second-panel rating, except one analyzed encounter that the second expert panel did not rate because its front video did not load for the reviewer. Comparisons with the second expert panel therefore include 407 encounters. Each reviewer viewed the front and side recordings with sound and recorded each criterion on a standard form. Because recordings were identified by reading number, reviewers were not masked to whether an encounter followed coaching. Second-panel reviewers did not see first-panel ratings, and no reviewer saw Coach-BP outputs. The second expert panel’s ratings were not used to train any model.

Two first-panel reviewers recorded only met criteria explicitly and left violated criteria blank. The third first-panel reviewer and all second-panel reviewers completed every cell explicitly. Blank cells were therefore interpreted as violations, and cells explicitly marked not available were excluded. The second expert panel confirmed 79–100% of the feet, back and legs violations recorded as blanks, but 8 of 16 and 28 of 49 of the arm violations recorded as blanks by the two first-panel reviewers who used that convention, against 16 of 17 recorded explicitly by the third. Agreement with the second expert panel does not depend on this interpretation. Coach-BP was evaluated separately against the two sets of expert ratings, without combining them, to assess whether performance was sensitive to the reference labels used for model development.

### 6. Statistical analysis and software

We estimated 95% CIs by percentile bootstrap with 2000 resamples of participants rather than encounters, so that encounters from the same participant were resampled together.^13^ Means across the five criteria took back supported from the side camera and were bootstrapped jointly, resampling participants once for all five criteria, and differences between two quantities measured on the same encounters were bootstrapped as paired differences. Agreement between Coach-BP and each clinician label set, and between the two sets of expert ratings, was quantified as balanced agreement, that is, balanced accuracy with the second expert ratings as the reference for every comparison with them, and as Cohen’s κ. The comparison between panels was used as a benchmark for agreement between two independently generated clinician reference sets, rather than as a formal estimate of within-panel inter-rater reliability.

Proportions are reported with Wilson score intervals, and sensitivity and specificity with exact Clopper– Pearson intervals. Tests were two-sided. Per-criterion and secondary comparisons were not adjusted for multiple testing and are interpreted descriptively. Missing landmark summaries were set to zero. Analyses used Python 3.11 with scikit-learn 1.8, NumPy 2.4, SciPy 1.17 and pandas 3.0, MediaPipe BlazePose 0.10.5, the DINOv2 ViT-B/14 checkpoint, the OWLv2 base patch-16 ensemble checkpoint and TalkNet weights pretrained on TalkSet with faces detected by S3FD, run with PyTorch 2.5 and Transformers 5.9. No performance target was prespecified, and accuracy is reported descriptively with 95% CIs. The analyses by reading, by agreement between the panels, by cuff side, and by camera tilt were exploratory. The out-of-fold score and verdict of every encounter for every criterion are stored with both expert panels’ ratings and are part of the data available on request.

### 7. Additional results

Sensitivity for detecting a violation ranged from 0·73 for the arm criterion to 0·93 for no talking (13 of 14 violations), and specificity from 0·83 for the arm criterion to 0·96 for legs uncrossed (Table A3).

**Table A3:** Cross-tabulation of Coach-BP against the expert ratings used in development.

| Criterion | TP | FN | FP | TN | No verdict | Sensitivity | Specificity |
| --- | --- | --- | --- | --- | --- | --- | --- |
| Feet flat | 56 | 16 | 36 | 300 | 0 | 0.78 (0.66–0.87) | 0.89 (0.85–0.92) |
| Arm criterion | 61 | 22 | 56 | 269 | 0 | 0.73 (0.63–0.83) | 0.83 (0.78–0.87) |
| Back supported | 43 | 4 | 33 | 326 | 2 | 0.91 (0.80–0.98) | 0.91 (0.87–0.94) |
| Legs uncrossed | 23 | 3 | 16 | 366 | 0 | 0.88 (0.70–0.98) | 0.96 (0.93–0.98) |
| No talking | 13 | 1 | 28 | 359 | 7 | 0.93 (0.66–1.00) | 0.93 (0.90–0.95) |

Violation is the positive class. Sensitivity and specificity are given with exact (Clopper–Pearson) intervals. Each row accounts for all 408 analyzed encounters, all of which had expert ratings for every criterion. No verdict counts encounters for which Coach-BP returned none (Section 1). Back supported is the test from the side camera. The test of the same model from the front camera is in Section 11 and Table A6. TP=true positive. FN=false negative. FP=false positive. TN=true negative.

Agreement with the second expert panel is given in Table A4 as balanced agreement and as Cohen’s κ, the usual measure of agreement between raters. When violations are rare, as here, κ depends strongly on how often each rater flags a violation, so raters who agree on most encounters can still have a low κ. Coach-BP flagged more violations than either panel for every criterion, which lowers its κ more than its balanced agreement. For no talking, for example, Coach-BP’s balanced agreement with the second panel was higher than the first panel’s (0·88 vs 0·84), but its κ was lower (0·38 vs 0·66). Across the five criteria, κ averaged 0·53 (95% CI 0·46–0·59) for Coach-BP and 0·77 (0·71–0·82) between the panels.

Considering the five criteria together, in the 401 encounters with all five verdicts, Coach-BP flagged at least one error in 136 of the 151 in which the first expert panel recorded one (sensitivity 0·90) and flagged none in 186 of the 250 without (specificity 0·74). All five verdicts were correct in 254 (63%) of the 401, and 123 (31%) had at least one false alert. Because almost half of the encounters were repeat readings after coaching, we also evaluated baseline readings alone. Mean balanced accuracy was then 0·86 and mean AUROC 0·91, with back supported from either camera. In encounters on which the two panels agreed, balanced accuracy was 0·86 for feet flat, 0·79 for the arm criterion, 0·95 for back supported from the side camera and 0·90 from the front camera, 0·92 for legs uncrossed, and 0·97 for no talking.

**Table A4:** Agreement with the second expert ratings (not used in development)

| Criterion | Coach-BP vs second ratings | Between both expert ratings | Difference (Coach-BP minus between ratings) | Cohen's $\kappa$, Coach-BP vs second ratings | Cohen's $\kappa$, between both expert ratings |
| --- | --- | --- | --- | --- | --- |
| Feet flat | 0.82 (0.76–0.87) | 0.90 (0.85–0.94) | –0.08 (–0.14 to –0.02) | 0.58 (0.48–0.68) | 0.81 (0.73–0.88) |
| Arm criterion | 0.75 (0.68–0.81) | 0.85 (0.80–0.90) | –0.10 (–0.18 to –0.03) | 0.38 (0.27–0.48) | 0.64 (0.53–0.74) |
| Back supported | 0.87 (0.81–0.92) | 0.87 (0.81–0.93) | –0.01 (–0.05 to 0.04) | 0.64 (0.51–0.74) | 0.80 (0.69–0.88) |
| Legs uncrossed | 0.92 (0.85–0.98) | 0.98 (0.93–1.00) | –0.06 (–0.14 to 0.01) | 0.67 (0.52–0.81) | 0.96 (0.88–1.00) |
| No talking | 0.88 (0.79–0.97) | 0.84 (0.71–0.95) | 0.04 (–0.04 to 0.18) | 0.38 (0.20–0.53) | 0.66 (0.39–0.84) |
| Mean of the five criteria | 0.85 (0.82–0.88) | 0.89 (0.85–0.92) | –0.041 (–0.074 to –0.004) | 0.53 (0.46–0.59) | 0.77 (0.71–0.82) |

Balanced agreement, with the second expert ratings as the reference, and Cohen's $\kappa$ . Differences are paired, on the same encounters, so they can show a difference even where the two intervals overlap. Comparisons use the 407 encounters with a second rating, less those without a Coach-BP verdict (Tables A1 and A3). Back supported is the test from the side camera, as in the main results. The test of the same model from the front camera is in Section 11 and Table A6. Intervals are 95% participant-bootstrap intervals.

Coach-BP matched or improved on the rule-based baselines for all five criteria, on the encounters scored by both, with clear gains in balanced accuracy of 0·29 (95% CI 0·21–0·36) for feet flat, 0·08 (0·01–0·14) for the arm criterion, 0·15 (0·08–0·22) for back supported from the side camera and 0·11 (0·03–0·20) from the front camera, and 0·14 (0·02–0·25) for no talking. The gain was 0·03 (−0·11 to 0·17) for legs uncrossed, and all five point estimates favored Coach-BP. Because the arm and feet baselines choose a variable and cut-off from the data, each was repeated with one encounter left out at a time. The arm baseline was stable (balanced accuracy 0·64–0·71), and the feet baseline was not (0·53–0·75). With its threshold chosen instead to maximize balanced accuracy on all the data, the no talking heuristic reached 0·84 and the difference from TalkNet was 0·09 (95% CI 0·03–0·16).

Arm AUROC was 0·90 (95% CI 0·85–0·94) with the cuff on the left arm and 0·76 (0·65–0·86) on the right, and agreement between the two expert panels was 0·88 and 0·77. Weighting the cuff sides equally in training, adding cuff side as an input and fitting a separate model for each side did not narrow the difference. Restricting the comparison to encounters whose detected and recorded cuff sides agreed left a similar difference (0·79 on the right against 0·91 on the left). One possible reason is that, with the camera on the cuffed side, more of the hand’s position fell along the line of sight. For the other criteria, AUROC differed by cuff side by at most 0·05, and every 95% CI for the difference in AUROC included zero.

The 38 encounters in which the detected arm differed from the staff record were examined in a further analysis, with one repetition of the cross-validation. In 35 of them staff recorded the left arm and the detector chose the right. They were filmed from further to the participant’s right (median 43° against 31°), and the detector was less certain in them. Using the staff-recorded arm did not improve accuracy. With the pose aligned to it, arm AUROC was 0·82 against 0·86 with the detected arm (difference −0·03 [95% CI −0·08 to 0·01]).

In a logistic regression including all four intake items, fitted to the 219 participants with complete responses, adjusted odds ratios for having at least one error ranged from 0·68 (95% CI 0·29–1·61) to 1·53 (0·65–3·59, global p=0·51). History of hypertension and antihypertensive treatment were concordant in 192 of 219 participants.

### 8. Model interpretation

The models’ inputs are landmark variables summarizing each recording, for example the mean sideways image position of a heel over all frames, its SD or its mean detection confidence. There were 363 for the arm criterion, 165 for legs uncrossed and 364 for feet flat. Several landmark variables often measure the same thing, so for interpretation they were combined into features. For each criterion, the Spearman correlation between every pair of landmark variables was computed across all encounters, and the variables were grouped by agglomerative hierarchical clustering with complete linkage on the distance 1 − |ρ|, cutting the tree at 0·3 so that every pair of variables within a feature had |ρ| of at least 0·7 (Figure S2). The outcome was not used. Cutting the tree at |ρ| of 0·65 or 0·75 instead gave the same 14 displayed features, with selection frequencies within 5 percentage points. Stability was assessed in 2000 refits of the L1 model, each on a random half of the participants.^14^ A feature was counted as selected in a refit if any of its landmark variables was retained, and features selected in at least 70% of refits that also had a standalone association with the criterion (single-feature AUROC of at least 0·55) are shown. Each feature’s combined measurement is the mean of its standardized landmark variables, with signs aligned. Its single-feature AUROC, the AUROC of that measurement used alone, is reported with a 95% CI from 2000 participant-bootstrap resamples. For back supported, whose scene features have no anatomical meaning, we instead edited the image and recomputed only the scene features, holding the pose features at their observed values. We grayed out either everything outside a box enclosing the patient and adjoining backrest or the box itself, setting the pixels to the mean color of the images DINOv2 was trained on, and compared each edit with graying out the same area at a random position. This was done for the side-view images and, separately, for the front-view images, with the model fitted in each fold on the unedited images of both views. The grouping into features was used only for interpretation. The models were trained and evaluated on the individual landmark variables, and none of these analyses affected model training or prediction.

Most features of the landmark models corresponded to what a clinician would assess (Figure 4A–C). For the arm criterion, the feature selected in every refit was the height of the cuffed hand above the hip midpoint (f1, single-feature AUROC 0·81 [95% CI 0·74–0·87]). It was followed by how far the cuffed hand sat in front of the hip midpoint (f3, selected in 89% of refits, AUROC 0·56 [0·49–0·64]) and how far the elbow and hand of the cuffed arm sat out to the side (f4, 87%, 0·74, and f5, 72%, 0·75), the pattern of an arm resting on a surface rather than hanging or lying in the lap. One arm feature, the detection confidence for the heel opposite the cuff (f2, 91%, 0·61), describes how clearly the lower body was seen rather than the arm itself. For legs uncrossed, the features were the sideways positions of the near and far foot and knee relative to the shoulder midpoint (f6, 97%, 0·63, and f7, 92%, 0·82), the height of the feet (f8, 89%, 0·73), and stillness of the near heel (f9, 79%, 0·93 [0·89–0·96]). For feet flat, they were the sideways position of the far foot (f10, 99%, 0·84 [0·79–0·89]), how far the near foot and knee sat in front of the hip midpoint (f11, 98%, 0·76), the output of the legs model (f12, 91%, 0·83), the height of the near foot (f13, 88%, 0·80), and the height of both feet (f14, 72%, 0·73). Near and far refer to the front camera. Because the legs and feet models were trained with mirrored copies, the same cues apply whichever side the camera is on.

For back supported from the side camera, graying out everything outside the box enclosing the patient and backrest left balanced accuracy at 0·90 (95% CI 0·84–0·94), against 0·94 for the unedited image in the same encounters, whereas graying out the same area at a random position reduced it to 0·72 (0·65– 0·78, Figure 4E, F). Graying out the patient box reduced balanced accuracy to 0·63 (0·56–0·69), whereas graying out the same area at a random position left it at 0·89 (0·83–0·94). The scene evidence therefore lies in the patient and chair rather than elsewhere in the room. These analyses used the 288 encounters, 31 with a violation, in which the pose model located the patient in every sampled side-view frame, as was needed to place the boxes. From the front camera the pattern was the same, in the 390 encounters, 45 with a violation, in which the patient was located in every sampled front-view frame. The values are in Figure 4E, F. On the unedited front-view images, balanced accuracy was 0·89. Figure S2 shows the composition of every feature, with each landmark variable’s own selection frequency.

### 9. Robustness to recording conditions

Robustness was tested without retraining. In each fold the model was fitted on unperturbed training participants, held fixed and applied to the held-out participant’s perturbed recording. We simulated a sideways shift of the frame of up to 10% of its width (about 15 cm at the patient), a zoom that changed apparent body size from 85% to 115%, and rotation of the frame by up to 10°. For the arm, legs and feet models the held-out participant’s landmark coordinates were transformed. For back supported and no talking, whose inputs are computed from the image itself, new inputs were generated by cropping, scaling and rotating real pixels of the original frames, with a central 80% crop as reference (75% for rotation) so that no content was invented at the image border. For back supported the fold model was fitted on the central 80% crops of the training participants’ recordings from both cameras, and it scored the held-out participant’s reframed recordings from each camera. For no talking, the full TalkNet pipeline, including face detection and tracking, was rerun on every encounter at every framing. Changes in camera viewpoint were simulated by rotating each held-out recording’s three-dimensional pose about the hips, round the patient (pan) or up and down (tilt), by up to 30° from the recording’s own camera angle, and projecting the change into the image coordinates. Within-recording movement variables were not rotated and pose was not re-estimated. Real and simulated viewpoints were grouped by absolute camera angle, measured as in Section 2. Robustness to recording quality was tested in the same way after the video was degraded (low light with sensor noise, reduced resolution, heavy compression or motion blur, each at three severities) or shortened to a random 75%, 50% or 25% of the recording, the mild, moderate and severe levels of recording length. A model was prespecified to fail a condition if AUROC fell by more than 0·03 or balanced accuracy by more than 0·05 with the paired bootstrap interval excluding zero. Augmentation was to be considered only for failures at mild or moderate severity, and failures only at the severe level were to be handled by recording-quality checks.

AUROC for arm supported, legs uncrossed and feet flat was unchanged by sideways shifts and changes in apparent size, and changed by at most 0·011 with rotation of the frame by up to 10° (Figure 3E–G). For back supported from the side camera, AUROC changed by at most 0·006 under shifts and size changes and by at most 0·007 with rotation. From the front camera it changed by at most 0·031 under shifts and size changes, and rotation by 10° in either direction failed the prespecified rule (Section 11). For no talking, AUROC was 0·975 at the reference crop and changed by at most 0·017 under shifts and size changes and by at most 0·010 with rotation. No face was tracked in 24 of the 6416 no-talking runs (401 encounters at 16 framings), and those encounters were left out of that framing only. At the operating threshold, balanced accuracy varied more than AUROC. Across shifts and size changes it ranged from 0·88 to 0·91 for back supported from the side camera, from 0·79 to 0·82 from the front camera, and from 0·89 to 0·93 for no talking, and across rotations of up to 10° from 0·76 to 0·79 for the arm criterion. Pooling the study’s own camera viewpoints with simulated ones by absolute angle, AUROC for legs uncrossed changed little with camera tilt from 10° upward to 50° downward (0·96–0·97) or with body angle from 20° to the patient’s left to 70° to the right (0·89–0·99). Feet flat was 0·89–0·91 with the camera 10° to 50° downward and 0·82–0·86 with the camera level or pointing up, and 0·90–0·93 across body angles. The arm criterion was 0·82–0·85 across tilt and 0·84–0·88 from head-on to 60°, falling to 0·73–0·81 with the camera on the patient’s left, to 0·78 at 70°, and to 0·69 at 80° (Figure S3A, B). In the study recordings alone, accuracy showed no consistent trend across thirds of the measured camera tilt (Figure S3C). No model failed at the mild or moderate level of any condition, and none failed under reduced resolution or motion blur at any severity. Failures are marked in Table A5. Severe low light (20% of normal exposure) degraded all four trained models, including back supported from either camera. No person was detected in 101 of the 406 side-camera recordings and 16 of the 408 front-camera recordings. Severe compression degraded legs uncrossed and back supported from either camera, and legs uncrossed also failed when only 25% of the recording was used.

**Appendix Figure S2:**
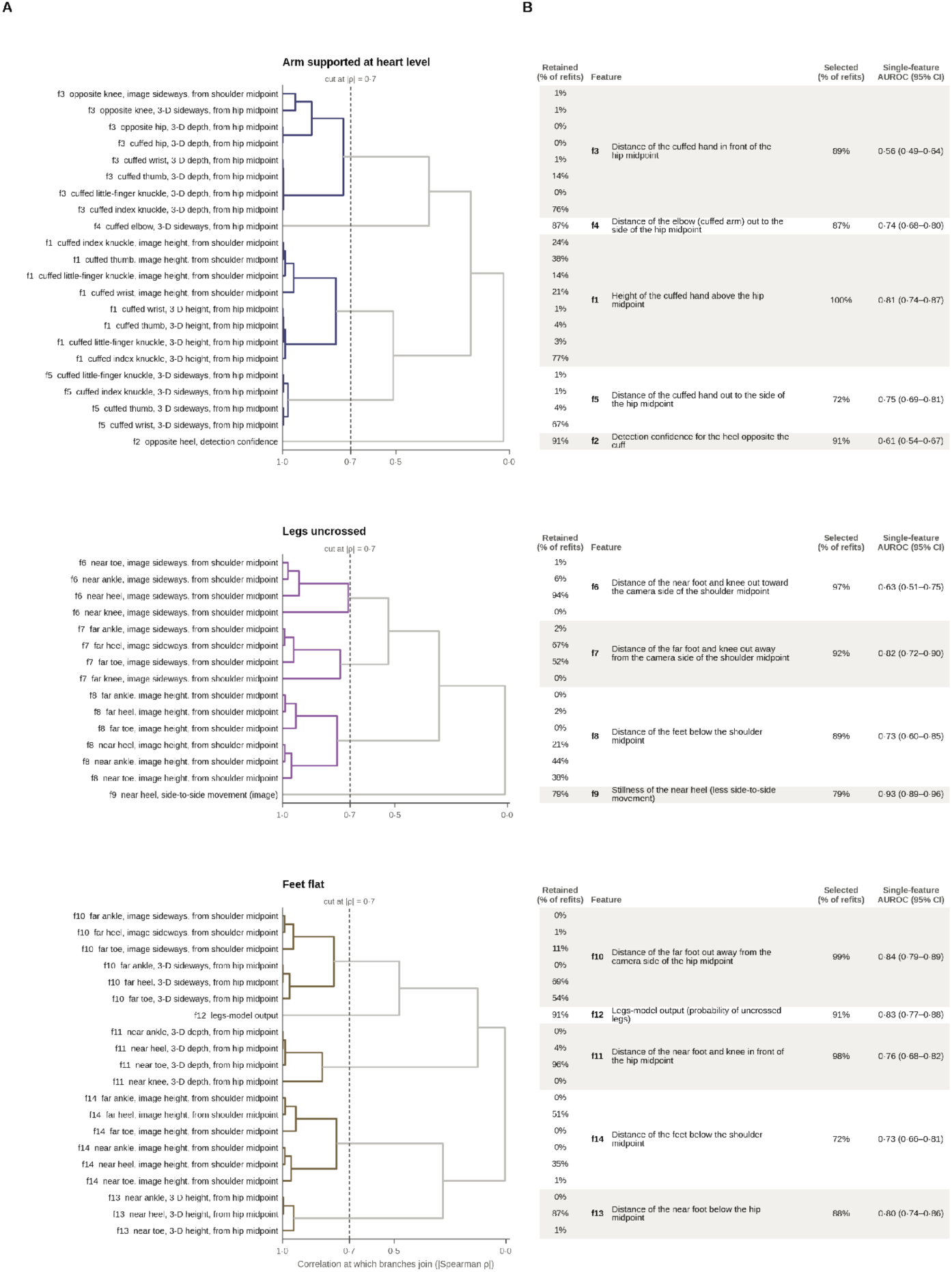
How the landmark variables were combined into the features of. Figure 4 (A) For each criterion, the dendrogram shows the hierarchical clustering (complete linkage on 1 − |Spearman ρ|) of the landmark variables that make up the displayed features. The horizontal axis gives the correlation at which branches join. Branches joined to the left of the dashed line (|ρ|=0·7) form one feature (colored, labeled f1–f14). The grouping is for interpretation only, and the models use each landmark variable separately. Image measurements are taken from the shoulder midpoint and divided by the shoulder-to-opposite-hip distance, and three-dimensional measurements are from the hip midpoint, in meters. Detection confidence is MediaPipe’s own score (0 to 1). Arm variables are named by cuff side (cuffed, opposite), because the arm pose is mirrored so that the cuffed arm is always on the same side. Legs and feet variables are named by the front camera (near, far), because those models were trained with mirrored copies, and the near side was the patient’s right in 95% of recordings. (B) Beside each variable, the percentage of 2000 refits on random halves of participants that retained it. Beside each feature, its name, the percentage of refits in which any of its variables was retained, and its single-feature AUROC with a 95% participant-bootstrap CI. Shading groups the rows of each feature. AUROC=area under the receiver operating characteristic curve.

**Appendix Figure S3:**
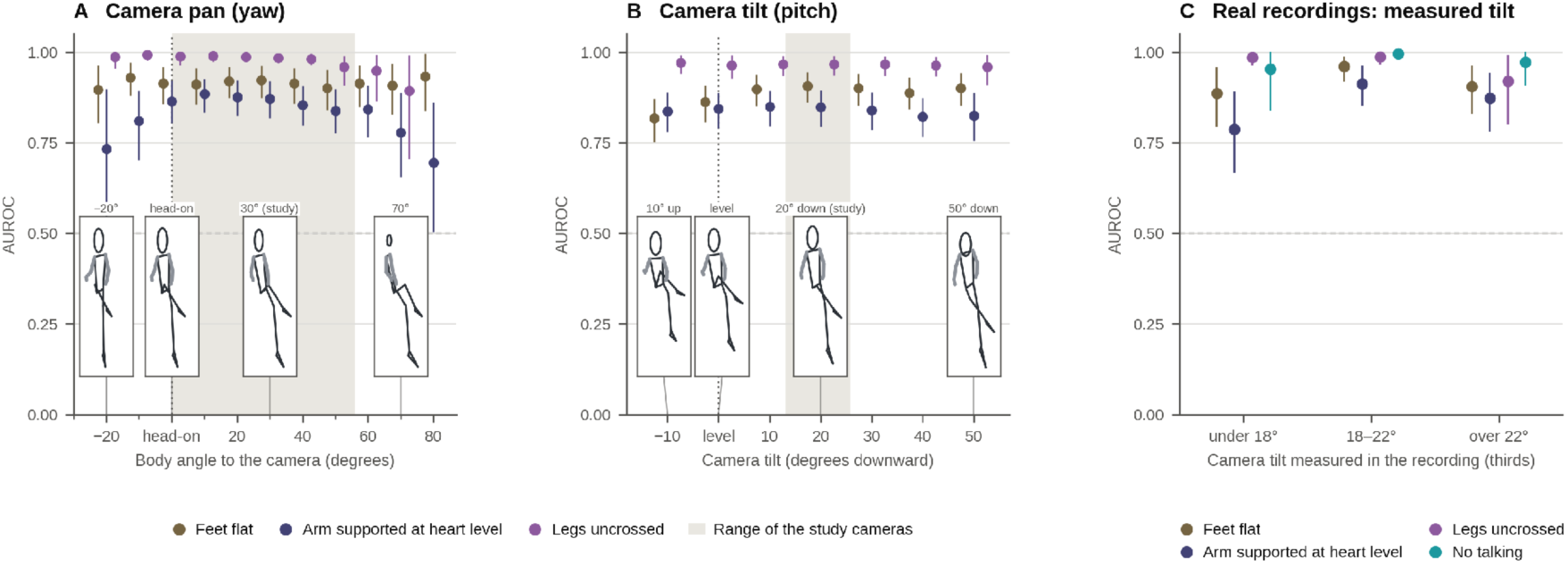
Robustness to camera viewpoint. (A, B) AUROC for arm supported at heart level, legs uncrossed, and feet flat by absolute camera angle. Panel A shows the body’s angle to the camera (pan, 0° head-on, positive with the camera to the patient’s right) and panel B the camera’s downward tilt (0° level, negative pointing up). Each held-out recording was re-angled in simulation by up to 30° from its own camera angle, by rotating its three-dimensional pose about the hips and projecting the change into the image. Results are grouped in 10° bins of the simulated camera angle, centered on multiples of 10° (level is −5° to 5°). The shaded band is the 5th–95th percentile of the study’s own cameras. Bins with fewer than 40 participants or 8 violations are not shown. Stick figures show the pose of a consented volunteer (not a study participant) re-angled in the same way, at the two ends of the range shown, at the reference view (head-on, level) and at the study cameras’ typical angle (30° pan, 20° down). (C) Real recordings only. AUROC for the four front-camera criteria within thirds of the camera tilt, measured in each recording from the room’s vertical edges (under 18°, 18– 22°, and over 22° downward, in the 388 recordings with at least ten vertical edges). Points show AUROC with 95% participant-clustered bootstrap intervals. AUROC=area under the receiver operating characteristic curve.

**Table A5:** Change in AUROC / balanced accuracy under degraded recordings, models held fixed.

| Condition | Arm criterion | Legs uncrossed | Feet flat | Back supported |
| --- | --- | --- | --- | --- |
| Low light, mild | -0.009 / -0.007 | -0.007 / -0.023 | -0.002 / -0.004 | -0.004 / 0.007 (2 no person) |
| Low light, moderate | -0.016 / -0.025 | -0.005 / -0.035 | 0.000 / -0.009 | -0.021 / -0.043 (17 no person) |
| Low light, severe | -0.099 / -0.080* (16 no person) | -0.136 / -0.183* (16 no person) | -0.135 / -0.143* (16 no person) | -0.152 / -0.152* (101 no person) |
| Resolution, mild | 0.001 / -0.008 | -0.008 / -0.013 | -0.002 / 0.001 | -0.001 / -0.012 (1 no person) |
| Resolution, moderate | -0.009 / 0.005 | -0.009 / -0.014 | -0.003 / 0.004 | -0.001 / -0.011 (1 no person) |
| Resolution, severe | -0.011 / -0.006 | -0.015 / -0.032 | -0.003 / -0.015 | -0.003 / -0.032 (1 no person) |
| Compression, mild | 0.003 / -0.006 | 0.002 / -0.021 | -0.004 / -0.007 | 0.002 / 0.006 (1 no person) |
| Compression, moderate | -0.002 / -0.011 | 0.005 / -0.001 | -0.003 / 0.020 | -0.005 / -0.037 (3 no person) |
| Compression, severe | 0.002 / 0.013 | -0.013 / -0.055* | -0.012 / -0.004 | -0.016 / -0.088* (2 no person) |
| Motion blur, mild | -0.003 / -0.009 | 0.001 / -0.014 | -0.004 / 0.001 | 0.001 / -0.011 |
| Motion blur, moderate | -0.004 / -0.009 | -0.009 / -0.013 | -0.003 / 0.004 | 0.002 / -0.013 (1 no person) |
| Motion blur, severe | -0.006 / 0.001 | -0.005 / -0.014 | -0.007 / -0.016 | 0.003 / -0.040 (1 no person) |
| 75% of recording | 0.003 / 0.000 | -0.018 / -0.054 | -0.011 / -0.004 | -0.001 / -0.004 |
| 50% of recording | 0.002 / 0.001 | -0.032 / -0.051 | -0.014 / -0.010 | -0.001 / -0.004 (2 no person) |
| 25% of recording | 0.003 / -0.005 | -0.048 / -0.072* | -0.022 / -0.020 | -0.001 / -0.010 (3 no person) |

\*Failure under the prespecified rule (AUROC down by more than 0.03 or balanced accuracy by more than 0.05, paired 95% interval excluding zero). Changes are relative to each model's unaltered recordings. Back supported is the test from the side camera of the one model for both cameras, and its fold models were fitted on the unaltered side and front recordings. Its results from the front camera are in the text.

### 10. Coaching

Of the 223 participants with an analyzed baseline reading, 171 also had an analyzed repeat reading after coaching. Because re-measurement was prompted by an observed error, this subgroup was enriched for baseline errors. Of the 171 re-measured participants, 128 (75%) had at least one error at baseline, compared with ten (19%) of the 52 without a repeat reading (odds ratio 12·5 [95% CI 5·8–27·0], p<0·0001). The paired comparison therefore describes change within a group selected for incorrect technique and does not represent a cohort-level change in prevalence. Among the 171 re-measured participants, the proportion with at least one technique error fell from 75% at baseline to 7% (12 participants) at the repeat reading, a difference of −67·8 percentage points (95% CI −74·8 to −60·8, p<0·0001). In all, 116 participants moved from at least one error to none. The mean number of errors per participant fell from 1·18 (SD 1·00) to 0·08 (0·33). All 43 participants whose baseline reading was already correct remained correct. With the second expert panel’s labels, whose raters had no role in coaching, the proportion with any error fell from 114 (67%) to 16 (9%) of the 170 participants with both readings rated by that panel. Of the 170 repeat readings with all five verdicts, Coach-BP flagged at least one error in 40 (24%), against 12 (7%) by the first expert panel. With back supported taken from the front camera it flagged 43 (25%).

Correction of an individual error was defined by the first-panel rating for each encounter. When an error was corrected between the first and last readings, Coach-BP classified the corrected reading as meeting the criterion in 22 of 22 cases for legs uncrossed, 50 of 53 (94%) for feet flat, 59 of 68 (87%) for the arm criterion, 32 of 35 (91%) for back supported from the side camera and 33 of 36 (92%) from the front camera, and nine of 11 for no talking.

### 11. One back supported model for both cameras

The study placed a second camera beside the patient because back support was expected to need a side view. To test this, one back supported model was trained on the side and front recordings of every training encounter together, with the same inputs from either view and nothing telling it which camera a recording came from. Each held-out participant’s side and front recordings were scored separately at the threshold of 0·5. The side test covers 406 encounters, 47 rated not supported, and is the main result. The front test covers 408, 48 rated not supported, and shows the same model from another camera position (Figure 3H).

From the side camera the model flagged 43 of the 47 encounters rated not supported, with 33 false alerts among the 359 rated supported. From the front camera it flagged 41 of 48, with 36 false alerts among 360 (Table A6). AUROC and balanced accuracy are in Table A6 and Figure 3H. On the 406 encounters with both recordings, the front camera differed from the side camera by −0·002 (95% CI −0·058 to 0·062) in AUROC and by −0·04 (−0·10 to 0·02) in balanced accuracy, and the two tests gave the same verdict in 348 (86%). Results were similar for first readings alone.

With back supported taken from the front camera, so that one camera served all five criteria, mean balanced accuracy across the five criteria was 0·87 (95% CI 0·84–0·90) and mean AUROC 0·93 (0·91– 0·94), against 0·88 (0·85–0·90) and 0·93 (0·90–0·94) with back supported from the side camera.

The expert ratings used in development rated back straight separately from back supported. From either camera the verdicts followed the rating of support much more closely than the rating of straightness (AUROC from the front camera 0·92 for not supported against 0·66 for not straight, difference 0·26 [95% CI 0·19–0·32]).

For comparison, two separate models were fitted with the same inputs on one view each (Table A6). On its own view each did about as well as the one model, and every paired difference included zero. The front-only model had the highest AUROC, 0·94 (0·91–0·97), but its scores were concentrated near 1, and at 0·5 it flagged 35 of the 48 encounters rated not supported. Neither carried over to the other view, with balanced accuracy 0·71 and 0·51.

Because the scores of the front-only model lay far from 0·5, a threshold set within each training fold was examined for every trained model, with a rule fixed before it was applied. In each fold the threshold maximized balanced accuracy on the training participants’ inner cross-validation scores and was applied to the held-out participant. For every model of the paper it stayed close to 0·5 (medians 0·47–0·55) and changed balanced accuracy by −0·019 to 0·009, with every interval including zero (Table A7). Only for the front-only model did it move far from 0·5, to a median of 0·77.

With the two-dimensional positions of the side recordings left as fractions of the frame, the AUROC, balanced accuracy, and agreement with the second expert ratings of the one model differed from those of the model as specified by at most 0·006 from either camera.

The robustness analyses of Section 9 were run for the one model from each camera. From the side camera no framing failed the prespecified rule. From the front camera sideways shifts and size changes passed, and rotation of the frame by 10° failed in either direction. AUROC fell by 0·04 (95% CI 0·01–0·08) at −10° and by 0·07 (0·02–0·12) at 10°, where balanced accuracy fell by 0·11 (0·04–0·18). On the central 80% of the frame, the model was as accurate as on the full frame from the side camera but not from the front camera (balanced accuracy 0·80 against 0·88, difference −0·07 [95% CI −0·13 to −0·02]).

These results show similar accuracy from either camera with one model, not that the cameras are equivalent. With 47 or 48 encounters rated not supported, the 95% CI for the difference in balanced accuracy reached 0·10 in favor of the side camera. The front camera was more sensitive to cropping and to rotation, so the result may depend more on how the camera is placed. The model was developed and tested at one site with one placement of each camera, and the choice to train it on both views followed a rule written before its results were seen, applied to cross-validated performance in this cohort (Section 4). The front camera was tested only at the position used in the study, and other positions could not be simulated for a model whose inputs include the image itself. It needs confirmation in recordings made elsewhere before a single camera is relied on for all five criteria.

### 12. Verdicts near the threshold

Verdicts were less accurate when the score lay near the threshold, so we examined withholding a verdict when the score lay between 0·3 and 0·7 for the four trained models, or when the maximum TalkNet score lay within 0·85 of its threshold of zero for no talking, which is the same distance on the logit scale (Table A8). This band was fixed before the results. A narrower band of 0·4–0·6, or a TalkNet score within 0·41 of zero, was added afterwards to show the trade-off, and no band is proposed as the one to use. With the band of 0·3–0·7, a verdict was given in 71%–92% of encounters across the criteria and mean balanced accuracy rose from 0·88 to 0·93, and with 0·4–0·6 a verdict was given in 86%–96% and the mean was 0·91. Each criterion withholds some verdicts, so all five verdicts were given in 125 (31%) of the 401 encounters at 0·3–0·7 and in 253 (63%) at 0·4–0·6. With the second expert ratings as the reference, the mean rose from 0·85 to 0·89 at 0·3–0·7, with no gain for no talking (0·88 either way).

At 0·3–0·7, an alert, a verdict that a criterion is not met, was right in 48%–78% of cases across the criteria, against 32%–61% with every verdict given, and AUROC among the verdicts given changed little (mean 0·93 with every verdict given, 0·93 at 0·4–0·6, and 0·94 at 0·3–0·7). The two expert panels did not clearly disagree more often on encounters inside the band (differences of −2 to 6 percentage points across the criteria, each 95% CI including zero). For no talking, with 14 encounters rated as talking, the one that the plain verdict missed has a verdict at 0·4–0·6 and none at 0·3–0·7, which moves balanced accuracy among the verdicts given from 0·94 to 0·98. Withholding a verdict was not part of the tool evaluated here, and its value for coaching is untested.

**Table A6:** Back supported by the recordings a model was trained on and the camera it was tested from.

| Trained on | Tested from | Encounters<br>(rated not<br>supported) | AUROC (95%<br>CI) | Balanced<br>accuracy (95%<br>CI) | Not<br>supported<br>flagged | Supported<br>passed |
| --- | --- | --- | --- | --- | --- | --- |
| Both views (the model of<br>the paper) | Side camera | 406 (47) | 0.92 (0.85–<br>0.98) | 0.91 (0.86–<br>0.95) | 43/47 (91%) | 326/359<br>(91%) |
| Both views (the model of<br>the paper) | Front camera | 408 (48) | 0.92 (0.87–<br>0.97) | 0.88 (0.82–<br>0.93) | 41/48 (85%) | 324/360<br>(90%) |
| Side recordings only | Side camera | 406 (47) | 0.92 (0.85–<br>0.97) | 0.90 (0.85–<br>0.95) | 41/47 (87%) | 334/359<br>(93%) |
| Side recordings only | Front camera | 408 (48) | 0.83 (0.77–<br>0.88) | 0.71 (0.64–<br>0.78) | 29/48 (60%) | 294/360<br>(82%) |
| Front recordings only | Front camera | 408 (48) | 0.94 (0.91–<br>0.97) | 0.85 (0.78–<br>0.90) | 35/48 (73%) | 346/360<br>(96%) |
| Front recordings only | Side camera | 406 (47) | 0.74 (0.66–<br>0.81) | 0.51 (0.48–<br>0.52) | 46/47 (98%) | 12/359 (3%) |

Each row is one test, with verdicts at 0.5. The model of the paper was trained on both views. The other two are separate comparison models, trained with the same inputs on one view and tested on both. Intervals are 95% participant-bootstrap intervals. AUROC=area under the receiver operating characteristic curve.

**Table A7:** Thresholds set within training folds.

| Criterion | Threshold set within training folds, median | Balanced accuracy at 0.5 | Balanced accuracy at that threshold | Difference (95% CI) |
| --- | --- | --- | --- | --- |
| Feet flat | 0.55 | 0.84 | 0.84 | 0.008 (−0.009 to 0.031) |
| Arm criterion | 0.55 | 0.78 | 0.79 | 0.004 (−0.019 to 0.031) |
| Legs uncrossed | 0.55 | 0.92 | 0.93 | 0.009 (−0.016 to 0.054) |
| Back supported from the side camera | 0.47 | 0.91 | 0.92 | 0.004 (−0.004 to 0.013) |
| Back supported from the front camera | 0.47 | 0.88 | 0.86 | −0.019 (−0.054 to 0.003) |

The median is taken over 2250 thresholds, one per held-out participant and repetition. Back supported is one model with one threshold in each fold, set from its side and front recordings together, so its two rows share a median. Differences are the threshold set within training folds minus 0.5 and are paired, on the same encounters, with 95% participant-bootstrap intervals.

**Table A8:** Accuracy when verdicts near the threshold are withheld.

| Criterion | Every verdict given: balanced accuracy (95% CI) | Withheld at 0.4–0.6: verdict given, n (%) | Withheld at 0.4–0.6: balanced accuracy (95% CI) | Withheld at 0.3–0.7: verdict given, n (%) | Withheld at 0.3–0.7: balanced accuracy (95% CI) |
| --- | --- | --- | --- | --- | --- |
| Feet flat | 0.84 (0.78–0.89) | 371 (91%) | 0.88 (0.82–0.92) | 304 (75%) | 0.90 (0.84–0.94) |
| Arm criterion | 0.78 (0.72–0.84) | 350 (86%) | 0.85 (0.79–0.90) | 291 (71%) | 0.87 (0.81–0.92) |
| Back supported | 0.91 (0.86–0.95) | 375 (92%) | 0.92 (0.87–0.96) | 328 (81%) | 0.93 (0.88–0.97) |
| Legs uncrossed | 0.92 (0.85–0.98) | 390 (96%) | 0.96 (0.90–0.99) | 375 (92%) | 0.96 (0.90–0.99) |
| No talking | 0.93 (0.85–0.97) | 356 (89%) | 0.94 (0.83–0.99) | 297 (74%) | 0.98 (0.97–0.99) |
| Mean of the five criteria | 0.88 (0.85–0.90) | – | 0.91 (0.88–0.93) | – | 0.93 (0.91–0.95) |
| Encounters with all five verdicts | 401 | 253 (63%) | – | 125 (31%) | – |
| All five correct among them | 254 (63%) | 191 (75%) | – | 108 (86%) | – |

The bands and their TalkNet equivalents are defined in Section 12. Percentages are of the encounters with a verdict for that criterion, and in the last row of the encounters in the row above. Back supported is the side-camera test, and the reference is the expert ratings used in development. Intervals are 95% participant-bootstrap intervals.

